# Transcriptome analysis in osteoarthritis primary tissues identifies high-confidence effector genes

**DOI:** 10.1101/2024.12.04.24318411

**Authors:** Georgia Katsoula, Ana Luiza Arruda, Mauro Tutino, Ene Reimann, Peter Kreitmaier, Karan M Shah, Diane Swift, Lorraine Southam, Siim Suutre, Galadriel Luzia Velazquez Silva, Kaspar Tootsi, Aare Märtson, Reedik Mägi, J Mark Wilkinson, Eleftheria Zeggini

## Abstract

Osteoarthritis, a whole-joint degenerative disorder, is a major public health burden that affects nearly 600 million individuals worldwide, but no disease-modifying treatment exists. Molecular profiling of relevant tissues is crucial for understanding the biological mechanisms underlying disease development. Here, we generate a comprehensive map of transcriptional regulation in disease-relevant primary tissues from knee osteoarthritis patients: macroscopically intact (low-grade, N=263) and degenerated (high-grade, N=216) cartilage, synovium (N=278), and fat pad (N=94). Of 9,738 unique expression quantitative trait loci (eQTL)-associated genes, 60.6% have not been reported previously. Using the largest osteoarthritis genome-wide association study (GWAS) to date, we find colocalization evidence with 117 genes, 67 of which had not been identified as effector genes before. We prioritise 38 high-confidence effector genes for osteoarthritis, based on multiple lines of molecular and functional genomics evidence, including *MUSTN1*, which is implicated in cartilage integrity. Our study provides insights into the molecular mechanisms underpinning osteoarthritis and offers much-needed drug repurposing opportunities.

## Introduction

Osteoarthritis, the most prevalent form of arthritis, is a chronic degenerative complex disease that is projected to affect over one billion individuals worldwide by 2050^1^. Osteoarthritis is characterized by progressive breakdown of articular cartilage and low-grade inflammation of the surrounding tissues^2,3^, leading to pain, stiffness, and reduced mobility^4^. It most commonly affects the knee, hand, and hip joints^5^. In addition to risk factors such as age, obesity, and joint injury, osteoarthritis also has a genetic risk component. Currently, there is no effective disease-modifying treatment for osteoarthritis, and strategies focus on pain management and joint replacement surgery. Therefore, there is an urgent need for an in-depth understanding of the processes underpinning osteoarthritis to foster the development of new therapies. Genome-wide association studies (GWAS) have identified over 960 independent risk variants for osteoarthritis^6^, the majority of which reside in non-coding sequences. The key challenge now is to identify the effector genes through which these associated variants affect osteoarthritis risk.

Osteoarthritis is a whole-joint disease, driven by a complex interplay of both shared and tissue-specific mechanisms^7^. Shared mechanisms involve inflammatory pathways, biomechanical stress and metabolic imbalances that collectively contribute to the degeneration of articular cartilage, the primary hallmark of osteoarthritis^4^. This includes extensive changes in the extracellular matrix (ECM), including loss of proteoglycans and collagen through enzymatic cleavage, mediated by metalloproteinases and aggrecanases and loss of chondrocyte ECM maintenance properties through senescence and apoptosis. In addition to cartilage degradation, osteoarthritis affects the synovium, subchondral bone and fat pad tissues. The synovium often exhibits inflammation (synovitis) characterized by increased production of pro-inflammatory cytokines, contributing to cartilage degradation and joint inflammation^2^. Pro-inflammatory cytokines are also secreted by the adipose tissue located in the infrapatellar fat pad^8^. There is evidence that crosstalk between the synovium and fat pad could stimulate the infiltration of immune cells and the secretion of catabolic molecules across both tissues^9^. Subchondral bone undergoes abnormal remodeling and sclerosis, influenced by altered bone turnover and microdamage repair processes^10^. These interconnected pathophysiological changes underscore the importance of studying multiple primary tissues.

Expression quantitative trait loci (eQTLs) capture the association between genetic variation and tissue-specific gene expression (proximal or distal)^11^. The regulatory relationships captured by eQTLs can reveal the function of disease-associated variants by identifying the genes and pathways they affect^12^. Given the tissue- and context-specificity underlying gene regulatory patterns^13^, eQTL regulatory relationships are best captured in the context of the studied phenotype in the primary affected tissue(s).

There have been two *cis-*eQTL studies in osteoarthritis primary tissues to date. Steinberg et al.^14^ identified *cis*-eQTLs in low-grade cartilage, high-grade cartilage, and synovium from up to 95 osteoarthritis patients of European ancestry. This study identified colocalization of eQTLs with osteoarthritis GWAS risk loci spanning five effector genes (*ALDH1A2*, *NPC1*, *SMAD3*, *FAM53A*, *SLC44A2*)^15^. A second study identified *cis-*eQTLs in the synovium of 202 Han Chinese individuals with osteoarthritis. The authors found evidence of colocalization with osteoarthritis GWAS loci for 34 genes with two, *FAM53A* and *SLC44A2*, overlapping with the European ancestry eQTL study^16^. These studies provide valuable insights into the proximal gene regulation mechanisms in osteoarthritis primary tissue. However, a well-powered eQTL map across multiple human joint tissues, including *trans-*acting variants, is still missing.

Here, we have generated *cis-* and *trans-*eQTL maps from knee joint primary tissues of 321 osteoarthritis patients of European ancestry, including macroscopically intact (low-grade) and degraded (high-grade) cartilage, synovium, and fat pad. This is the first eQTL study for fat pad tissue and the sample size for the other three tissues is increased substantially compared to previous studies (low-grade cartilage: 3.02 times, high-grade cartilage: 2.25, synovium: 1.38 times). We identify shared and disease tissue-specific biological mechanisms involved in osteoarthritis and integrate this resource of gene expression molecular data with knee-related osteoarthritis GWAS to gain insights into loci associated with disease risk.

## Results

### Osteoarthritis primary tissues *cis-*eQTL discovery

We performed *cis*-eQTL analyses in 321 knee osteoarthritis patients (Figure 1, Table S1), achieving a significant increase in statistical power across the allele frequency spectrum (Figure S1) while also replicating previous findings (Figure S2). We identify 6,646 *cis-*eGenes (genes targeted by at least one eQTL) in low-grade cartilage and 4,639 in high-grade cartilage, including 8,378 and 5,355 conditionally independent *cis*-eQTLs, respectively (Figure 2, Table S2). In synovium we find 6,411 *cis-*eGenes targeted by 8,043 conditionally independent *cis*-eQTLs (Figure 2, Table S2), and in fat pad osteoarthritis tissue, we identify 592 *cis-*eGenes regulated by 606 conditionally independent *cis-*eQTLs (Figure 2, Table S2).

**Figure 1:**
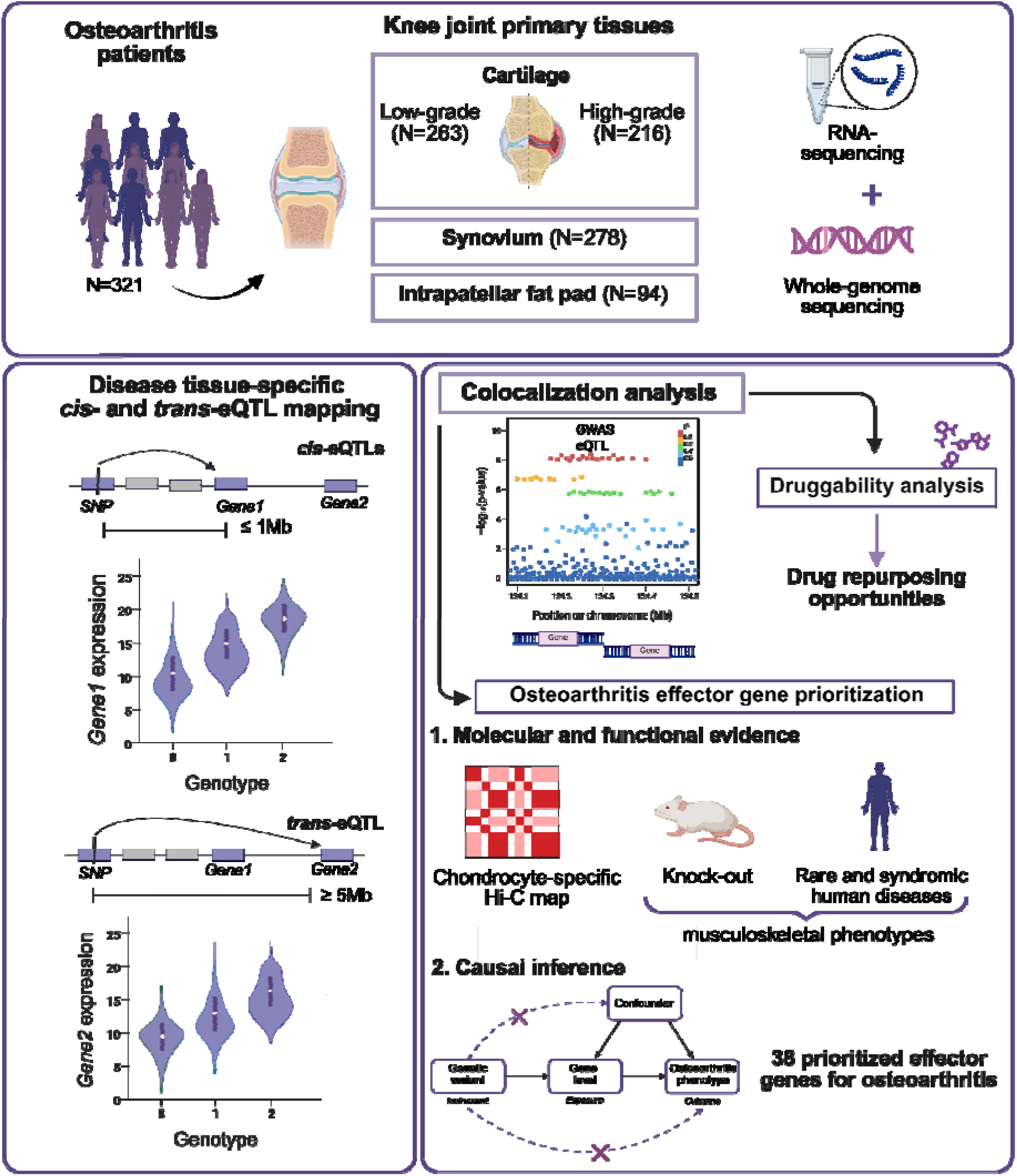
Study design.

**Figure 2:**
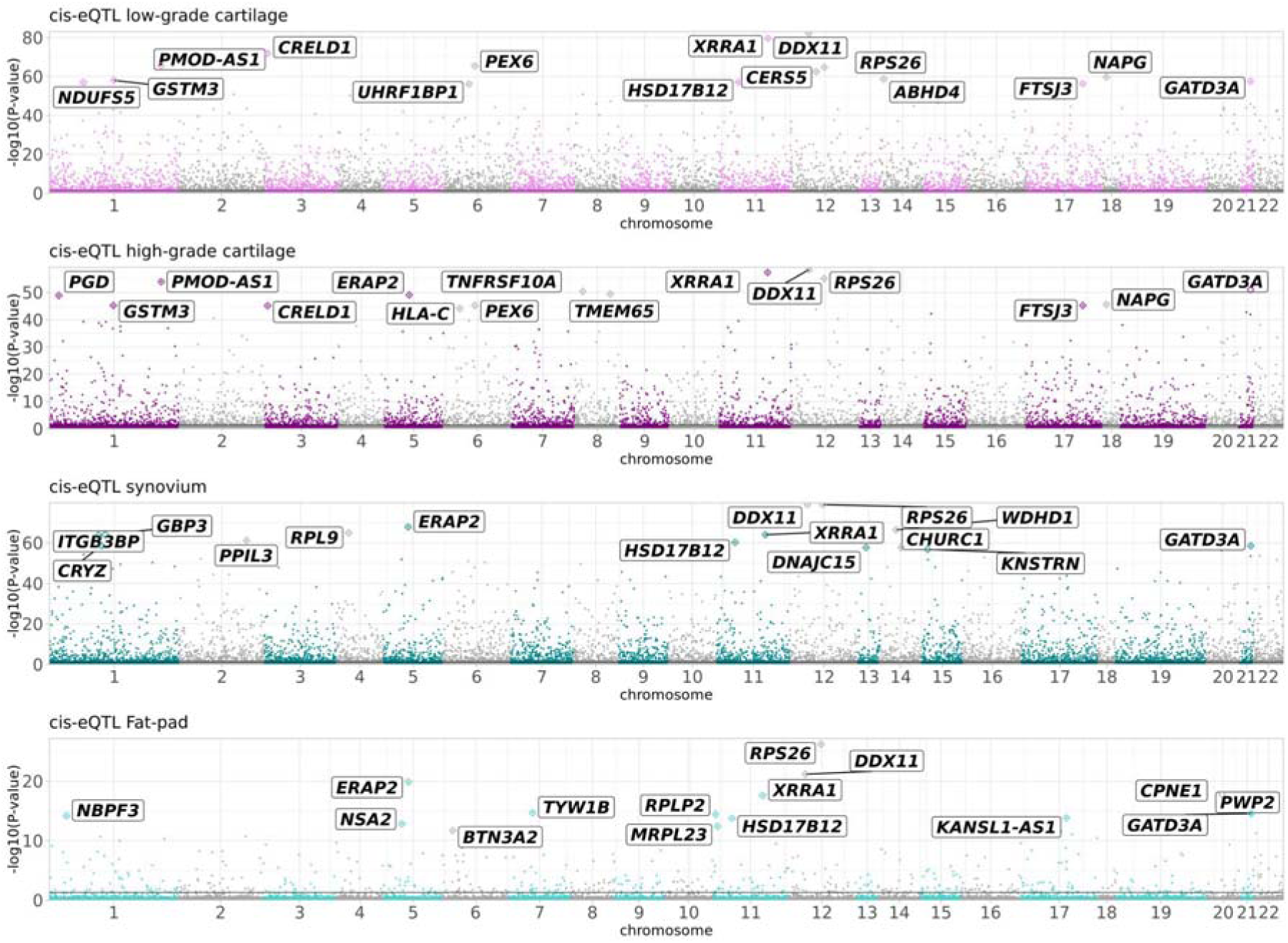
Genomic distribution of *cis-*eQTLs. Manhattan plots depicting the negative log of the permutation p-value (q-value) of the most significant association per gene across all variants within 1 Mb of the gene’s transcription start site for low-grade cartilage, high-grade cartilage, synovium and fat pad tissues. Top 15 *cis-*eGenes per disease tissue are labeled in black.

### Distinct eQTL effects in osteoarthritis tissues

Understanding how genetic regulation varies across osteoarthritis-affected tissues is important for deciphering the specificity of disease mechanisms. We sought to identify distinct genetic regulatory effects present in individual disease tissues using two complementary approaches. Firstly, we investigated eQTLs regulating genes expressed exclusively in a single disease tissue and not in others, identifying 597 unique eGenes in cartilage, 70 in synovium, and 14 in fat pad (Table S3). Secondly, to formally assess distinct genetic regulatory patterns across osteoarthritis tissues for commonly expressed genes, we performed a meta-analysis across all examined disease tissues (low-grade cartilage, high-grade cartilage, synovium and fat pad), and identify 53 additional eGenes with eQTL effects exclusive to low-grade cartilage, 24 to high-grade cartilage, 267 to synovium, and 54 to fat pad, with no effect in other tissues (Figure S3, Tables S4-S5).

Several genes showed large eQTL effects in a single disease tissue and are related to osteoarthritis biology. *CXCL14*, a chemokine associated with osteoarthritis pain^17^ and regulating connective tissue during limb development^18^, shows distinct regulation of its expression in low-grade cartilage (*CXCL14:*rs35039539) (Figure S3, Table S4). *CXCL14* also acts as a natural inhibitor of the *CXCL12/CXCR4* axis, which is known to regulate chondrocyte hypertrophy^19^ and is a potential therapeutic target for osteoarthritis^20^.

*SNAI1* (*SNAI1*:rs437276) and *CXCL5 (CXCL5:*rs17230459) show distinct expression regulation in high-grade cartilage (Figure S3, Table S4). *SNAI1* is a transcription factor involved in chondrocyte proliferation through repression of *COL2AI* and *ACAN* transcription *in vitro*^21^ and has also been shown to act downstream of FGFR3 signaling in mouse chondrocytes, regulating both Stat and MAPK and ERK1/2 pathways. The latter are critical for chondrocyte terminal differentiation^22–24^.

In the synovium, we find distinct regulation for *C1QTNF1*, *DMRT2* and *RAMP3*, among other genes (Figure S3, Tables S4-S5). *C1QTNF1* codes for an adipokine that enables collagen binding, has a role in inflammation, and is involved in chondrocyte maturation with activation of the ERK1/2 signaling pathway^25^. *DMRT2* codes for a transcription factor involved in skeletal development and endochondral bone formation^26^, while *RAMP3* is involved in angiogenesis, which also takes place in osteoarthritis and is linked to pain and synovitis^27^. Several other genes with proposed roles in osteoarthritis show specific regulatory patterns in synovium including *TIMP3*, a critical inhibitor of matrix metalloproteinases (MMPs), *GSK3B* inducing osteoarthritis through activation of Wnt pathway in cartilage^28^ and *SOX6* a transcription factor with established roles in synovial joint development^29^.

*AMPH*, coding for a protein involved in synaptic vesicle endocytosis^30^, shows the most distinct regulation in osteoarthritis fat pad compared to other tissues (Figure S3, Tables S4-S5), pointing to a possible role in local neuroinflammatory processes. Several further genes with fat pad– specific regulation, have been connected to osteoarthritis. For instance, *IL34* is involved in macrophage differentiation and is associated with synovitis, supporting its role in local immune activation^31^. *PER1*, a core circadian clock gene, with a proposed role in suppressing chondrocyte differentiation^32^, has been linked to the temporal regulation of inflammatory mediators^33^, which may contribute to daily variation in osteoarthritis symptoms. *ZEB2* is the mediator of the differentiation of age-associated B cells^34^, which may contribute to the low-grade inflammation observed in osteoarthritis Together, these genes underscore the unique immunoregulatory and neuroimmune environment of the osteoarthritis fat pad.

### Shared eQTL effects among osteoarthritis primary tissues

In line with GTEx findings^35^, we observe widespread sharing of eQTL effects across osteoarthritis-relevant tissues, reflecting common regulatory architecture. Specifically, 72.3% of *cis*-eQTLs are shared across all four tissues when considering both effect direction and magnitude (see Methods). Tissue pairs with closer biological similarity show even greater overlap, with 85.9% of *cis*-eQTLs shared between low- and high-grade cartilage, and 73.6% between synovium and fat pad (Figure 3A).

**Figure 3:**
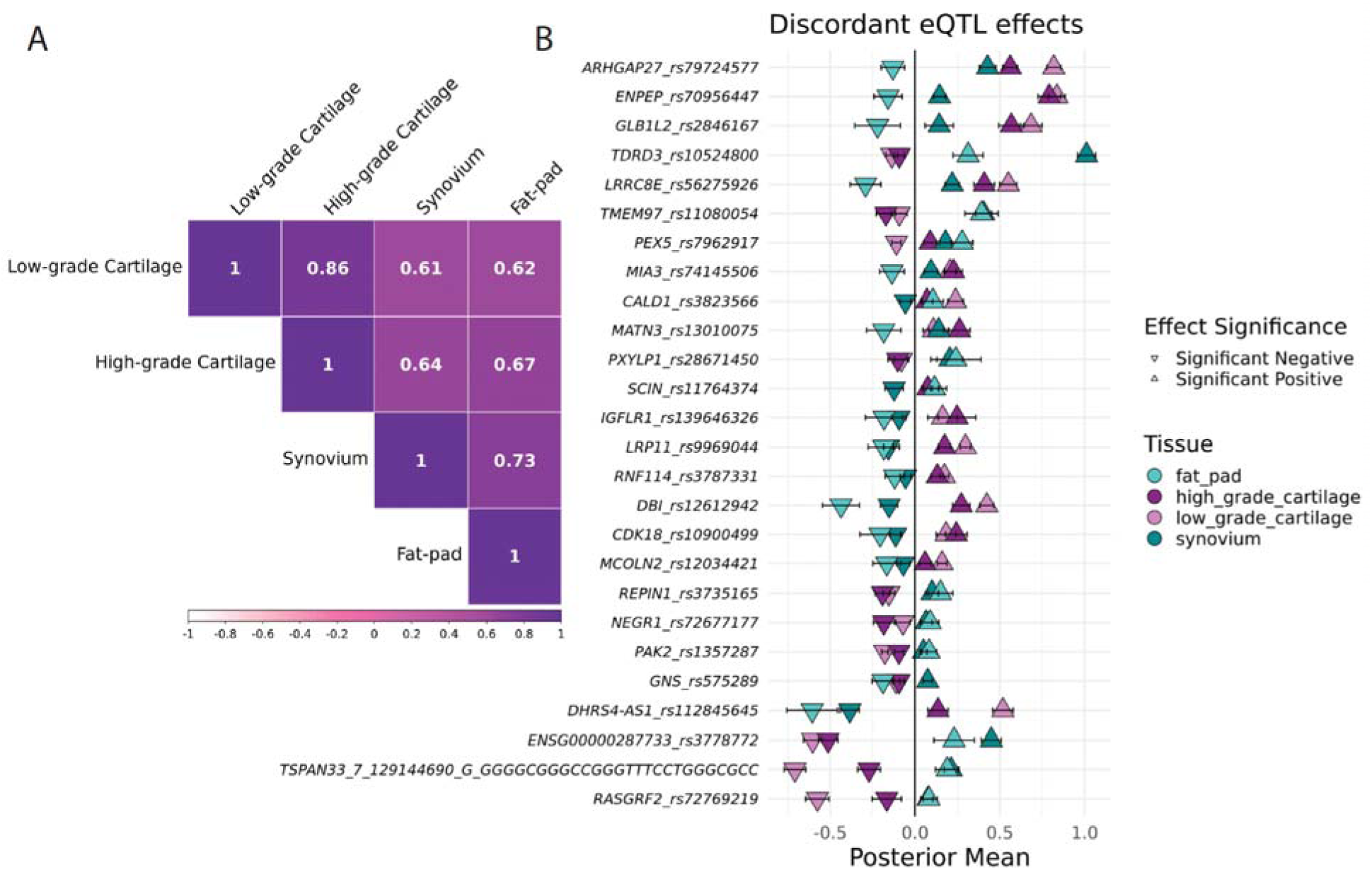
Patterns of shared and discordant eQTL effects among osteoarthritis primary tissues. (A) Heatmap showing pairwise sharing by magnitude of *cis-*eQTLs among tissues. (B) 26 eQTL–eGene pairs showing opposite directions of effect between osteoarthritis tissues.

Nevertheless, a subset of 26 eQTL–eGene pairs, though significant across all disease-relevant tissues, exhibit discordant effects (Tables S4-S5), showing opposite directions of association in at least one tissue (Figure 3B). In 14 out of 26 cases, the direction of effect was concordant between low- and high-grade cartilage, as well as between synovium and fat pad, reflecting greater similarity within these tissue pairs. However, we also identified cases with opposite effects in a single tissue. For example, the insertion allele (TGCAC) of the rs70956447 variant (reference allele T), associated with *ENPEP* expression, shows a positive effect on *ENPEP* expression in cartilage (both low- and high-grade) and synovium, but a negative effect in the expression of the same gene in the fat pad. *ENPEP* encodes glutamyl aminopeptidase, an enzyme involved in angiogenesis and also a marker of common skeletal progenitors in the postnatal limb bone^36^. Among the genes with opposite effects specifically in fat pad, we observed several involved in ECM remodeling and previously linked to osteoarthritis, including *MATN3* and *MIA3* (Figure 3B). These discordant eQTLs may reflect tissue-specific regulatory mechanisms or context-dependent gene regulation relevant to osteoarthritis pathophysiology.

### Disease progression eQTLs in osteoarthritis cartilage

Cartilage was the only tissue that included both low- and high-grade osteoarthritis samples, enabling the investigation of stage-specific regulatory changes that may reflect disease progression. We identified 117 eQTL–eGene pairs with effects exclusive to low-grade cartilage and 38 pairs with effects specific to high-grade cartilage (Figure S4, Tables S5-S6).

Among the genes with low-grade-specific regulation, several are implicated in pathways central to chondrocyte function and cartilage integrity. These include *PIK3R1*, *MTOR*, *INSR*, and *PREX1*, key components of the PI3K/AKT/mTOR signaling axis, which regulates chondrocyte survival, metabolism, and autophagy^37^. We also observed regulation of *PTGER2*, a prostaglandin receptor linked to cartilage inflammation^38^, and *ADAMTS5*, a central aggrecanase in osteoarthritis pathophysiology^39^. Additional low-grade-specific effects were found for *AHR*, which plays an important role in the immune system within the skeletal niche and the differentiation of mesenchymal stem cells into other cell lineages including chondrocytes and adipocytes^40^ and *FOXJ1*, a ciliogenesis regulator with emerging roles in chondrocyte mechanotransduction^41^. These findings highlight that regulatory variation in early-stage osteoarthritis preferentially affects genes involved in signaling, inflammation, and matrix turnover in cartilage.

In high-grade cartilage, several eQTLs were associated with genes involved in oxidative stress (*MT1F*, *OSGIN2*), inflammation (*CXCL5*, *SELE*), and DNA repair (*RAD50*, *NUP98*). *TMEM165*, important for glycosaminoglycan chain elongation and chondrocyte maturation^42^, also showed high-grade-specific regulation. Additionally, we identified lncRNAs *ALDH1A3-AS1* and *LINC01614* as targets of high-grade-specific eQTLs. *ALDH1A3-AS1* may influence retinoic acid signaling via its sense gene *ALDH1A3*, which is associated with higher expression of chondrocyte-specific genes such as *COL2A1* and *SOX9*^43^. *LINC01614*, previously reported to be upregulated in lesioned versus preserved osteoarthritis cartilage^44,45^, may contribute to disease-related transcriptional changes. Together, these findings suggest that regulatory variation in advanced-stage osteoarthritis increasingly impacts genes involved in cellular stress responses, matrix regulation and transcriptional reprogramming.

Notably, we identified four eQTL–eGene pairs that exhibited significant associations in both low- and high-grade cartilage, but with opposite directions of effect (Figure 4A, Table S7). These included *TTPAL*, *MAF1*, *SPATA13*, and *TMEM220* (Figure 4B). Such directionally discordant effects may reflect dynamic, stage-specific regulatory mechanisms, potentially altering gene function or cellular context as osteoarthritis advances.

**Figure 4:**
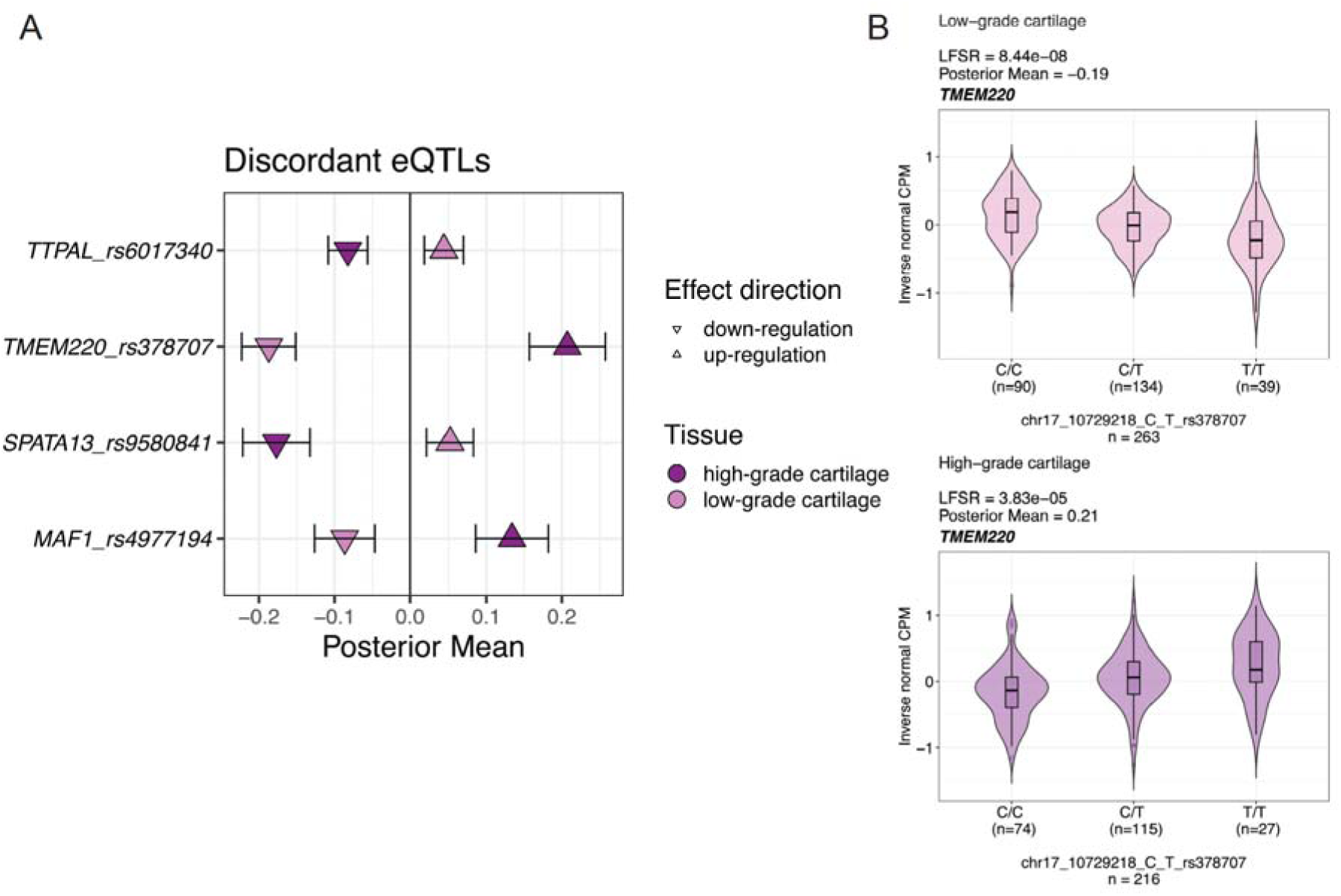
Disease progression eQTLs. (A) Four eQTL–eGene pairs exhibit opposite directions of effect between low- and high-grade osteoarthritis cartilage, indicating stage-specific regulatory changes. (B) Violin plot illustrating the rs378707 C allele, which is associated with decreased *TMEM220* expression in low-grade cartilage but increased expression in high-grade cartilage.

### *Trans*-eQTLs is osteoarthritis primary tissues

We investigated distal gene regulation (≥5 Mb from transcription start sites) in osteoarthritis primary tissues. In low-grade cartilage, we identified 363 *trans*-eQTLs (12 independent signals) regulating 12 *trans*-eGenes; in high-grade cartilage, 180 *trans*-eQTLs (5 independent) for 5 genes; in synovium, 163 *trans-*eQTLs (6 independent) for 5 genes; and a single *trans*-eQTL– eGene pair in fat pad. Four *trans*-eGenes - *UQCRHL*, *ORC6*, *MYLK3*, and *KANSL1-AS1* - were shared between low- and high-grade cartilage, with *KANSL1-AS1* also appearing in fat pad and *UQCRHL* in synovium (Table S8, Figures S5-S6).

Most *trans*-eGenes were also *cis*-regulated: 9/12 in low-grade cartilage, 4/5 in high-grade, and 2/5 in synovium. These included genes linked to mitochondrial function and metabolism (*UQCRHL*, *TEFM*, *COQ8A*), cell cycle and DNA replication (*ORC6*, *CDK10*), and protein degradation (*DCUN1D5*, *TRIM69*). Notably, *CRHR1* and *KANSL1-AS1*, both located in the osteoarthritis-associated 17q21.31 locus, show elevated expression in knee osteoarthritis cartilage compared to non-osteoarthritis controls^46^.

To explore potential mechanisms underlying these distal regulatory effects, we next asked whether *trans*-eQTL variants might influence gene expression by disrupting transcription factor binding sites^47^. Across all tissues, we identified 46 unique variants predicted to affect motifs for 57 transcription factors (Table S9), with several linked to retinoic acid signaling, including RXRA, RXRG, RARA::RXRA, RXR::RAR_DR5, RORA, and RORC. In synovium, multiple variants associated with *TRIM69* expression are predicted to disrupt retinoic acid-related binding sites, suggesting that retinoic acid-responsive regulatory elements may mediate these effects. Although direct regulation of *TRIM69* by these factors has not been shown, other TRIM family members, such as *TRIM32*, interact with retinoic acid receptors^48^. As retinoic acid signaling is a pathway previously implicated and proposed as a therapeutic target in osteoarthritis^6,52^ its *trans*-regulation may be of particular relevance for the disease.

### Resolution of osteoarthritis GWAS signals

We conducted statistical colocalization analysis between knee-related osteoarthritis GWAS risk loci^6^ and primary knee osteoarthritis tissues *cis*-eQTLs (Table S10, Table S11). Among the 211 osteoarthritis GWAS risk loci examined, 74 (34.1%) showed evidence of a shared association signal with *cis*-eQTLs from primary osteoarthritis tissues, spanning 117 unique genes. 67 out of the 117 colocalizing genes have not been previously prioritized as effector genes for osteoarthritis^6^.

To glean biological insights from the 117 colocalizing genes, we performed biological pathway over-representation analysis (Table S12). Compared to all eGenes across all four primary osteoarthritis tissues, the colocalizing genes are enriched for osteoblast differentiation (q-value=0.037) as well as signaling pathways including transforming growth factor beta (TGF-β) (q-value=0.037), mitogen-activated protein kinase (MAPK) (q-value=0.042) and phosphoinositide 3-kinase/protein kinase B (PI3K/AKT) (q-value=0.048) (Figure 5). These three signaling pathways have been previously implicated in osteoarthritis^37,49,50^, as well as osteoblast differentiation^51^. The 117 colocalizing genes are also enriched for gene ontology pathways from broad biological mechanisms including cell fate and morphogenesis, as well as for 9-cis-retinoic acid (9cRA) metabolic process (q-value=0.032). While all-trans retinoic acid has been more extensively studied in osteoarthritis^52^, 9cRA may have similar effects due to its ability to bind to retinoic acid receptors and influence retinoid signaling.

**Figure 5:**
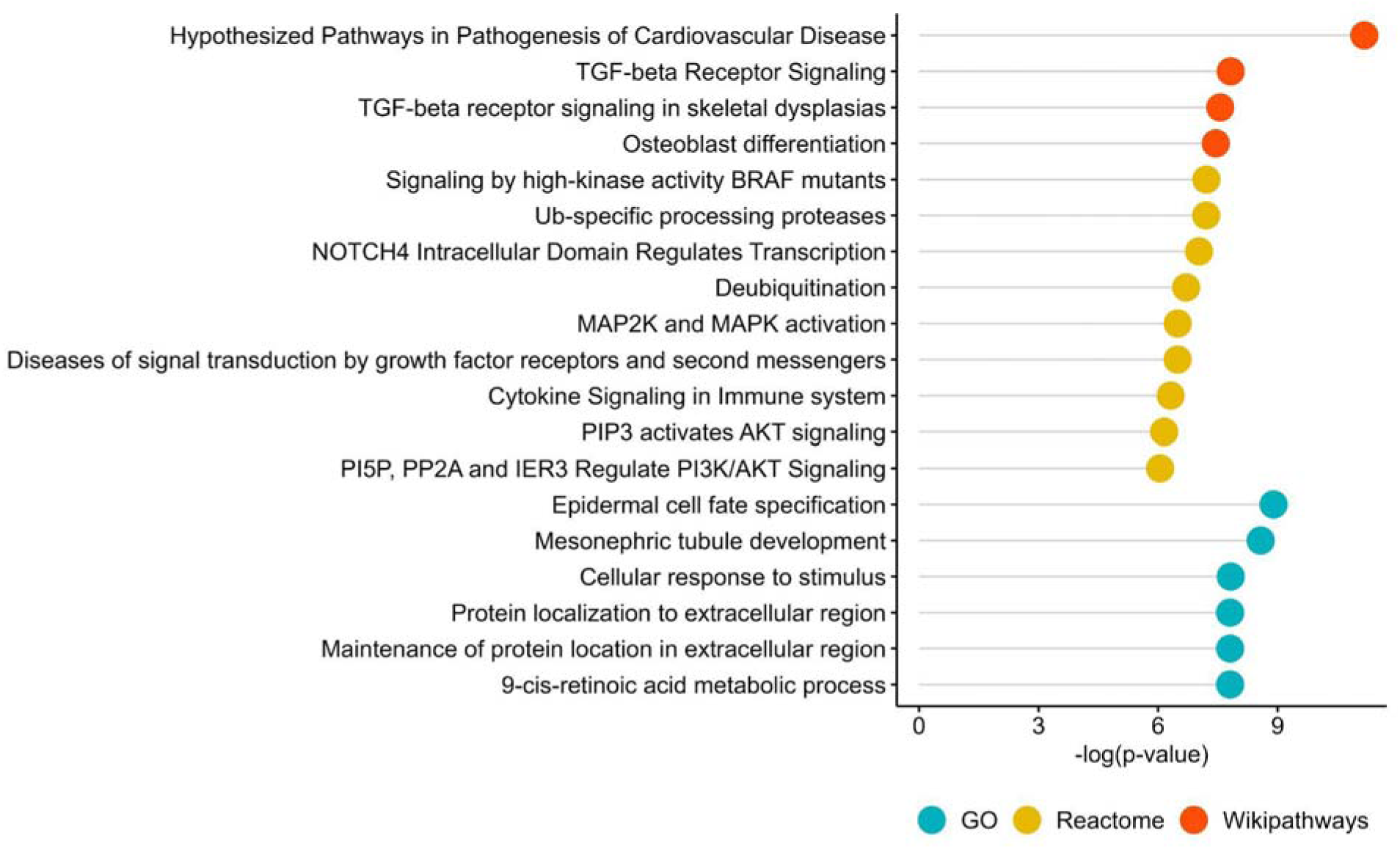
Pathway enrichment analysis of the colocalizing genes. Results (q-value<0.05) of pathway-based over-representation analysis using ConsensusPathDB^53^ on the 117 genes colocalizing with knee-related osteoarthritis phenotypes. All eGenes across all four primary osteoarthritis tissues studied here were used as the background.

For each colocalizing signal, we calculated a 95% credible set of variants, within which the shared causal variant lies with 95% probability. We studied the location of these variants by overlaying them with active enhancer-promoter loops from primary knee osteoarthritis chondrocytes^54^. Given that the high-throughput chromosome conformation capture (HiC) data were derived from cartilage tissue, we focused primarily on colocalizing regions in high-grade and low-grade cartilage. Two eQTLs prioritized by the colocalization analysis are located within enhancers of active enhancer-promoter loops for the same respective eGene: rs10948 (*SLC44A2*) and rs12932078 (*WWP2*). This suggests that these eQTL-eGene pairs may exert their effect on knee osteoarthritis by altering enhancer-promoter interactions. *WWP2* and *SLC44A2* have been previously identified as osteoarthritis effector genes^6^ (Figure 6, Figures S7-8).

**Figure 6:**
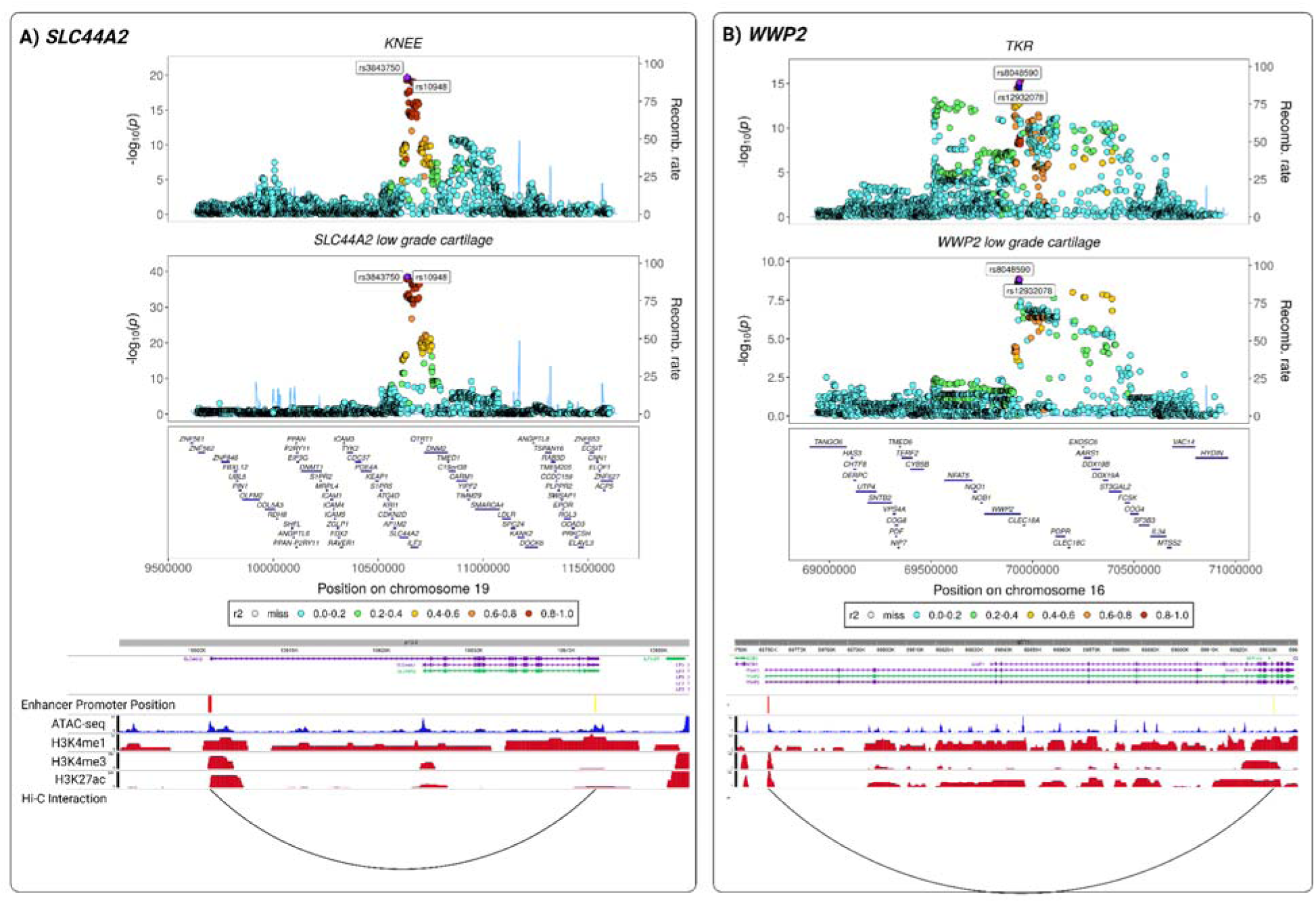
Examples of colocalizing *cis*-eQTLs in low-grade cartilage overlapping with chondrocyte active enhancer-promoter loops derived from Hi-C data. (A) Example of rs10948, a colocalizing *cis*-eQTL of *SLC44A2* in low-grade cartilage and knee osteoarthritis found in the enhancer region of an active enhancer-promoter loop. (B) Example of rs12932078, a colocalizing *cis*-eQTL of *WWP2* in low-grade cartilage and total knee replacement found in the enhancer region of an active enhancer-promoter loop. For both examples: Upper panel: Regional association plot depicting the highlighted colocalizing region. The enhancer-overlapping *cis*-eQTL and lead colocalizing SNP are labeled. Lower panel: Track plot generated using WashU Epigenome Browser^55^, showing annotated gene transcripts, SNP positions (yellow), enhancer and promoter locations (red), chromatin states, and histone modifications derived from chondrocyte ATAC-seq^56^ and ChIP-seq^57^ data (TKR= total knee replacement, KNEE = knee osteoarthritis).

Three cartilage eQTLs colocalizing with osteoarthritis GWAS signals reside within active promoters: rs3742566 (*LGALS3*), rs2102066 (*WWP2*) and rs35346340 (*FES*) (Figures S9-11). This suggests that these loci may play a critical role in the proximal regulation of gene expression and contribute to knee osteoarthritis risk by influencing promoter activity. Additionally, three colocalizing synovium eQTLs reside within active promoters: rs12129705 (*ZNF697*), rs11539637 (*FES*) and rs117827273 (*MSL1*) (Figures S11-13). *MSL1* and *ZNF697* have not been prioritized as effector genes in the latest osteoarthritis GWAS meta-analysis^6^.

### Prioritization of 38 high-confidence effector genes

To infer putative causal effects of the identified colocalizing eGenes on knee-related osteoarthritis phenotypes, we conducted tissue-specific causal inference analyses using Mendelian randomization (MR)^58^ (Table S13). Out of the 117 colocalizing genes, 87 show evidence of a causal effect on knee-related osteoarthritis for matching disease phenotype and tissue^59^. In addition to causal inference evidence, we have scored the genes based on orthogonal lines of evidence including knockout mouse phenotypes^60^, rare and syndromic human disease phenotypes^61^ and HiC information in cartilage^54^ (Table S14). Out of the 117 colocalizing genes 40 are associated with musculoskeletal-related phenotypes (see Methods) in knockout mouse models and 10 in rare and syndromic human diseases. We prioritize 38 genes that showed at least two lines of evidence pointing to their involvement in osteoarthritis, in addition to colocalizing with GWAS signals (Figure 7). Of these, 12 have not been defined as osteoarthritis effector genes in the latest GWAS meta-analysis^6^.

**Figure 7:**
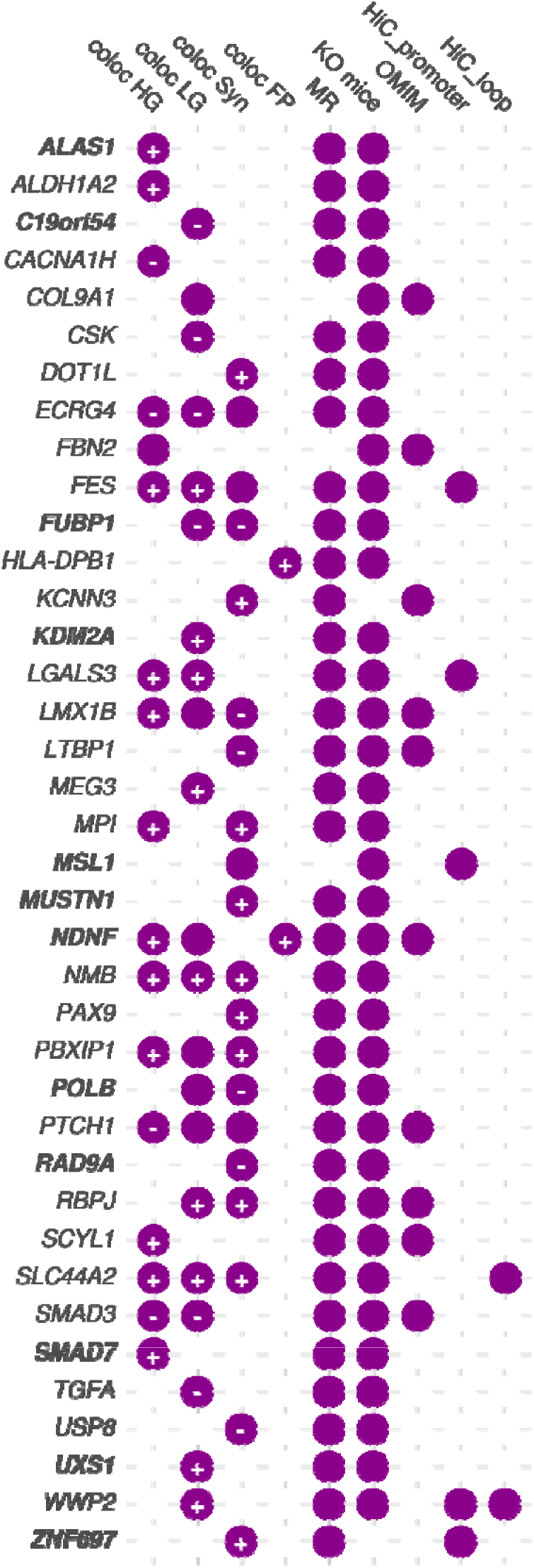
Prioritized colocalizing genes based on additional evidence of involvement in knee-related osteoarthritis. In addition to showing the osteoarthritis primary tissue of colocalization (HG=high-grade cartilage, LG=low-grade cartilage, Syn=synovium, FP=fat pad), the heat map visualizes the lines of evidence used to assess involvement in disease, including evidence of knockout mouse models (KO mice), rare and syndromic diseases (OMIM), causal inference analysis (MR), overlap with active promoter regions from chondrocytes (HiC_promoter) and overlap with enhancer-promoter loops from chondrocyte (Hi_loop). The signs (+/-) in the colocalization columns indicate the direction of putative causal effect identified by MR. The genes highlighted in bold indicate genes that were not previously prioritized as effector genes for osteoarthritis in the latest disease GWAS meta-analysis^6^.

We further evaluated the spatial expression patterns of the colocalizing genes using spatial transcriptomics data from osteoarthritis degraded cartilage (see Methods). We find that 37 out of the 38 prioritized colocalizing genes are expressed in at least one of the analyzed cartilage zones (Table S15). All of these genes are detected in the superficial layer of osteoarthritis cartilage. Nine genes (*ALDH1A2, FBN2, LMX1B*, *LTBP1, MSL1*, *POLB*, *SCYL1*, *SMAD7* and *UXS1*) are also expressed in the middle zone, while three genes (*ALDH1A2, HLA-DPB1* and *RBPJ*) are expressed in the deep cartilage zone. This shows that the majority of prioritized genes are active in the superficial layer of osteoarthritis cartilage, potentially reflecting its increased involvement in osteoarthritis pathology.

Among the prioritized effector genes, *WWP2*, an established effector gene for osteoarthritis^6^, showed the strongest evidence of involvement, with four lines of evidence (Figure 7). Variants associated with the expression of this gene colocalize with knee osteoarthritis (PP4=0.98) and knee replacement surgery (PP4=0.89) in low-grade cartilage. One of the variants in the 95% credible set of the colocalization signal, rs2102066 resides in a chondrocyte active promoter-enhancer loop, suggesting a potential mechanism of action of this signal and *WWP2* on osteoarthritis. We find evidence that increased expression of *WWP2* in low-grade cartilage is causally associated with an increased risk of knee osteoarthritis (OR=1.23, q-value=6.34×10^−38^) and knee replacement surgery (OR=1.37, q-value=2.84×10^−42^), whereas increased expression in synovium has a protective effect on the risk of knee osteoarthritis Our findings point to contrary mechanisms of action for *WWP2* in cartilage and synovium. We find evidence of expression of *WWP2* in the superficial zone of osteoarthritis cartilage.

*NDNF* accrued three lines of evidence in support of its role in osteoarthritis and has not been prioritized as an effector gene before. This gene colocalizes with knee osteoarthritis in low-grade (PP4=0.79, PP4/PP3=3.86) and high-grade cartilage (PP4=0.75, PP4/PP3=3.03) as well as in fat pad tissue (PP4=0.76, PP4/PP3=3.31). We find evidence of a risk-increasing effect of *NDNF* expression on knee osteoarthritis risk in high-grade cartilage (OR=1.1, q-value=6.69×10^− 17^) and fat pad (OR=1.05, q-value=5.54×10^−13^). *NDNF* also showed evidence of expression in the superficial layer of osteoarthritis cartilage. Knockout mice for *NDNF* show skeletal muscle atrophy. Mutations in this gene are linked to the rare human disease hypogonadotropic hypogonadism 25 with anosmia, which is characterized by delayed or absent puberty and is associated with delayed bone age. *NDNF* promotes neuron migration, growth and survival, and may also be involved in regulating growth factor activity^62^. Neurotrophic factors can modulate inflammation and pain perception, an important symptom for osteoarthritis^63,64^.

### Prioritized genes are enriched for chondrocyte-related pathways

The 38 prioritized colocalizing genes are enriched for gene ontology pathways related to morphogenesis, cell fate, cell signaling, cell regulation and metabolism (Table S16). Some genes are involved in osteoarthritis-relevant pathways specifically associated with cartilage, chondrocyte phenotypic stability and bone (Table S16). For instance, *MUSTN1* and *SMAD7*, two genes that were not previously prioritized as effector genes for osteoarthritis^6^, are involved in the chondrocyte proliferation (q-value=0.012) and cartilage development pathways (q-value=0.035). *SMAD7* is further involved in regulation of ossification (q-value=0.003).

*SMAD7* encodes an intracellular inhibitory SMAD protein that acts as a negative regulator of the TGF-β signaling pathway. TGF-β promotes chondrocyte proliferation and extracellular matrix synthesis, preserving cartilage integrity^65^. Variants associated with the expression of *SMAD7* in high-grade cartilage colocalize with total knee replacement (PP4=0.81) and show evidence of a putative risk-increasing causal effect on total knee replacement risk (OR=1.19, q-value=3.47×10^−5^). *SMAD7* is the target of mongersen sodium, a SMAD-7 mRNA antisense inhibitor in clinical trials for Crohn’s disease and ulcerative colitis. By inhibiting SMAD7, mongersen sodium aims to restore TGF-β activity, pointing to a potential repurposing opportunity of this drug for osteoarthritis. Knockout mouse models for *SMAD7* present abnormal bone marrow cell physiology^66^, which can be associated with bone marrow lesions common at late stage osteoarthritis.

Variants associated with the expression of *MUSTN1* in synovium colocalize with knee osteoarthritis GWAS (PP4=0.86). In the same tissue, we find evidence of a risk-increasing effect of *MUSTN1* expression on knee osteoarthritis (OR=1.11, q-value=6.18×10^−16^). *MUSTN1* has been implicated in critical processes for maintaining cartilage integrity and function, such as chondrogenesis, chondrocyte differentiation, and proliferation^67^. The protein encoded by *MUSTN1* is predominantly expressed in skeletal muscle and bone and plays a role in myoblast differentiation, bone regeneration, chondrogenesis, and muscle hypertrophy^68^ . Knockout mouse models for this gene show skeletal muscle fibrosis^69^.

### Translational drug repurposing opportunities for the colocalizing genes

Considering the translational potential of drug targets with genetic evidence supporting their effect on disease^70^, we conducted a druggability search on the 117 colocalizing genes using the Open Targets database^71^. A total of 14/117 genes are linked to drugs with at least phase II clinical trials completed (Table S17). *TGFA* and *CACNA1H* are the only colocalizing genes targeted by a drug with phase II clinical trials for osteoarthritis completed. *TGFA* is the target of fepixnebart, an antibody that inhibits TGFA interaction with its receptor, thereby hindering downstream signaling pathways. In addition to osteoarthritis, fepixnebart is under clinical trial with phase II completed for neuropathic pain and low back pain. Pregabalin is an anticonvulsant drug used to treat neuropathic pain conditions and fibromyalgia that targets *CACNA1H.* This drug is used as chronic pain management in the treatment of osteoarthritis symptoms. *CACNA1H* is the target of several additional drugs, all of which act as an antagonist or blocker.

Two further genes targeted by drugs in completed phase II clinical trials are associated with osteoarthritis: *FGF1* and *MAPK3*. *FGF1* has been associated with osteoarthritis through its role in cartilage degradation and repair mechanisms. Studies suggest that *FGF1* may contribute to cartilage repair by promoting chondrocyte proliferation and extracellular matrix synthesis while also modulating inflammation^72,73^. Drugs targeting *FGF1* span inhibitory, agonistic and activator mechanisms of action. Variants associated with the expression of this gene colocalize with knee osteoarthritis, osteoarthritis at any site and total knee replacement in low-grade osteoarthritis cartilage (Table S10, Table S11).

*MAPK3* is implicated in osteoarthritis by influencing chondrocyte function and inflammatory responses. Studies have shown that MAPK3 regulates chondrocyte proliferation, apoptosis, and the production of matrix metalloproteinases, enzymes that degrade cartilage matrix, thereby contributing to osteoarthritis progression^74^. It has been shown that MAPK inhibitors are protective against early stage osteoarthritis^75^. Both drugs targeting *MAPK3*, ulixertinib and temuterkib, are inhibitors of this gene. In addition to *MAPK3* eQTLs colocalizing with knee-related osteoarthritis in synovium (PP4=0.69, PP4/PP3=9.62), we find evidence of a protective effect of *MAPK3* expression on knee osteoarthritis in the same tissue (OR=0.87, q-value=7.21×10^−5^).

Six further colocalizing genes targeted by drugs with at least phase II clinical trials completed have not been previously prioritized for osteoarthritis: *ALAS1*, *PSMA4, TNFRSF9, FRK, PIK3R3* and *KCNK3* (Table S17). *KCNK3* encodes a member of the two-pore domain potassium channel family, which is the target of sevoflurane. Sevoflurane, a widely used inhaled anesthetic, has been shown to modulate the activity of various ion channels and is indicated for pain treatment.

## Discussion

Large efforts have been made to identify genetic risk signals for osteoarthritis. However, elucidating the function and tissue of action of these variants is challenging. Here, we have generated the largest *cis*- and *trans*-eQTL maps in low-grade cartilage (n=263), high-grade cartilage (n=216), synovium (n=278) and fat pad (n=94) knee osteoarthritis joint tissues. The availability of this resource aids in uncovering shared and disease tissue-and stage-specific biological mechanisms. By combining the newly-generated eQTL maps with osteoarthritis GWAS and HiC data of patient chondrocytes, we prioritize effector genes and highlight the mechanism of action of risk variants. By focusing on primary tissues directly affected by osteoarthritis, the identified *cis-*eQTLs provide a more precise and context-specific understanding of the genetic architecture underlying the disease highlighting the distinct tissue-specific genetic regulatory mechanisms operating in osteoarthritis-affected tissues.

Comparing our results with the first *cis-*eQTL study in cartilage and synovium^14^, we recapitulate ∼70% of *cis-*eQTL-gene pairs across tissues with largely concordant effect sizes. The comparison of our study with the synovium *cis-*eQTL map in 201 osteoarthritis patients of Han Chinese ancestry^16^ yielded a 50% overlap for eGenes. This may be attributed to population-specific effects, including differences in allele frequencies and linkage disequilibrium patterns, and/or to analysis approach differences.

Colocalization between the *cis*-eQTL maps and knee-related osteoarthritis GWAS finds evidence of a shared signal for 34.1% of the tested loci, yielding 117 unique colocalizing genes. The partial elucidation of GWAS signals by colocalization with eQTL data underscores the increasing evidence that eQTLs and GWAS identify partially different sets of associated variants partly due to natural selection^76^. We prioritized 38 colocalizing genes supported by multiple lines of evidence of involvement in osteoarthritis. All were causally linked to knee-related osteoarthritis risk. Considering that genetic evidence support can accelerate drug discovery^77^, the prioritized genes represent appealing candidates for clinical translation, including drug repurposing opportunities.

Despite the gain in power to elucidate GWAS signals presented in this study, the generated data are restricted to samples from individuals of European ancestry. This reflects a well-recognized gap in the availability of molecular data from global populations^78^. Going forward, it is important that efforts focus on closing this gap. Recent single-cell RNA-seq (scRNA-seq) studies in osteoarthritis cartilage^79^, synovium^80^ and fat pad^81^ have implicated specific cell types involved in osteoarthritis. Additionally, *trans-*eQTLs have been shown to capture cell-type specific effects^82^. Combining genomic data with scRNA-seq in larger sample sizes is needed to differentiate between real regulatory effects and different cell compositions.

Robust molecular data from primary tissues can be valuable in enhancing the translation of genetic findings for all complex diseases. Here, we have generated the largest molecular eQTL maps for four osteoarthritis-relevant tissues and two cartilage disease stages. Leveraging these powerful data, we detect shared and tissue-specific biological mechanisms underlying knee osteoarthritis. Our findings identify new candidate effector genes for this disease along with tissue of action and direction of effect, pointing to drug repurposing opportunities.

## Methods

### Sample selection

In this study, we included osteoarthritis patients who had undergone total knee replacement surgery for osteoarthritis with no history of significant knee surgery, infection, fracture, or malignancy within the previous 5 years. Additionally, we ensured that no patient had been treated with corticosteroids (systemic or intra-articular) within the previous 6 months, or any other drug associated with immune modulation. We isolated matched cartilage samples from weight-bearing regions of the joint along with synovium and fat pad tissue from the same patients. Cartilage samples were also scored macroscopically using the International Cartilage Repair Society (ICRS) scoring system^83^. We collected from each patient one sample of ICRS grade 0 or 1, representing low-grade osteoarthritis degradation, and one sample of ICRS grade 3 or 4, representing high-grade osteoarthritis degradation. All study participants provided informed consent and samples were collected under Human Tissue Authority license 12182 and National Research Ethics Service approval 15/SC/0132 and 20/SC/0144, South Yorkshire and North Derbyshire Musculoskeletal Biobank, University of Sheffield, UK.

### Library preparation

#### RNA-sequencing

We extracted RNA using Qiagen AllPrep RNA Mini Kit according to the manufacturer’s instructions and as previously described in Steinberg et.al^14^. We isolated and sequenced samples in five batches that were later uniformly processed. In brief, we isolated Poly-A-tailed RNA from total RNA using Illumina’s TruSeq RNA Sample Prep v2 for batches 1-4, while for the 5th batch, we used SMART-Seq® Ultra® Low Input RNA Kit. The libraries were then sequenced on the Illumina HiSeq 2000 and HiSeq 4000 (75bp paired-ends) (batches 1-4) and Novaseq6000 (batch 5) recovering a median of 80.8 million reads per sample.

#### Whole-genome sequencing (WGS)

WGS data were generated from patients’ DNA as previously described^84^. In brief, libraries were prepared from DNA samples using the standard Illumina paired-end DNA protocol as per manufacturers’ instructions. Then, they were amplified and sequenced in two batches on the NovaSeq platform.

### Preprocessing and quality control (QC) of RNA-sequencing data

We performed alignment of the fastq files to the reference genome Ensembl GRCh38 release 105 (cDNA and non-coding) using STAR version 2.7.9a.^85^ We summarized the reads using FeatureCounts (Subread 2.0.3) software and called duplicates using PicardTools [https://broadinstitute.github.io/picard/]. We further used RSeqQC (version 4.0.0)^86^ to summarize alignment metrics. We first performed sample-level quality control and excluded samples that did not pass the following criteria: RNA integrity number >5, percentage of uniquely mapped reads >70%, reads mapping to genomic features >40%, reads mapping to intronic regions or rRNA <30%, reads that failed RseQC strandness check <30% and including >10 million reads. Using the aforementioned criteria we excluded 42 low-grade cartilage samples, 35 high-grade cartilage samples, 14 synovium samples, and 13 fat pad adipocyte samples. This way we ensured the inclusion of only high-quality samples.

We further excluded outlier samples in a tissue-specific manner. Outliers were identified using robust principal component analysis considering the first three principal components (PcaGrid method rrcov R package)^87^. We excluded samples from individuals that had bilateral knee replacement where low- and high-grade disease cartilage samples were pooled (7 individuals). For individuals with bilateral knee replacement, we excluded one pair of matched cartilage samples each (3 samples), keeping only the sample pair with the best quality. For samples with technical replicates we also kept to one with the best quality (2 samples). The final number of samples after the exclusions across tissue was 881, derived from 331 patients (low-grade cartilage:272, high-grade cartilage:223, synovium:288, fat pad:98). After QC we performed principal component analysis (PCA) to find main drivers of variation and evaluate the presence of batch effects (Figure S14). The principal component 1 (PC1), explaining 29.89% of the variation in the data, corresponded to the different tissues, while PC2, explaining 13.22% of the variation, corresponded to the sequencing batches.

### Preprocessing and QC of Whole Genome Sequencing (WGS) data

WGS preprocessing and QC was performed as previously described^84^. CRAM files were converted to BAM format using samtools (version samtools conda version 1.14). The later were further converted to fastq format using *bedtools bamtofastq* function (bedtools conda version 2.30.0). We performed variant calling per batch and using Sarek nf-core pipeline (version 2.7.1, https://nf-co.re/sarek/2.7.1) with the additional options “*--tools HaplotypeCaller, -- generate_gvcf*”. We further performed joint variant calling by adapting the GATK4 pipeline (https://github.com/IARCbioinfo/gatk4-GenotypeGVCFs-nf) to be used with GATK (docker container broadinstitute/gatk:4.2.5.0).^88^ For all variant calling purposes GRCh38 reference was specified.

For variant-level quality control, we used Variant Quality Score Recalibration (VQSR) method specifying a tranche threshold of 99.5% for SNPs and 99% for INDELS. The expected false positive rate for SNPs was 2.5% and the expected sensitivity was 97%.

We excluded samples with high heterozygosity rate (2 samples), non-reference allele concordance with the directly typed genotype data using variants MAF >L0.01 (1 sample - also showing heterozygosity rate), outliers in sequencing depth (1 sample), sex mismatched (2 samples), relatedness > 0.2 (2 samples), ethnicity outliers (2 samples). Ethnicity outliers (3 individuals) were identified using the Ancestry and kinship toolkit (based on 1000G data from phase three; https://github.com/Illumina/akt/tree/master)^89^. Nine samples were excluded in total.

After sample-level QC, we performed additional filtering and excluded variants with MAF <L0.01, Hardy-Weinberg equilibrium PL<L10^−5^ and call rate <L=L0.99. After selecting individuals matching WGS and RNA-seq data per tissue we performed further variant filtering to keep only bi-allelic and common variants (MAF > 0.05). The final WGS data sets per tissue after filtering were as follows: low-grade cartilage (263 individuals, 6,304,167 variants), high-grade cartilage (216 individuals, 6,323,956 variants), synovium: (278 individuals, 6,338,438 variants), fat pad (94 individuals, 6,295,898 variants).

To characterize population structure and ensure consistency across datasets, we next performed principal component analysis (PCA) on the filtered genotype data using bcftools^90^, PLINK^91^, and the 1000 Genomes Project reference panel^89^. Genotype data in VCF format were filtered to retain only unique samples across all tissues using *bcftools view* and updating (MAF ≥ 0.05), call rate (CR ≥ 99%), and biallelic status as described above. The resulting VCF contained 7.285.082 variants across 321 samples. The resulting VCF was indexed and converted to PLINK binary format. To assess population structure, we downloaded phased genotype data from the 1000 Genomes Project (GRCh38), normalized and converted it to PLINK format, and LD-pruned each chromosome. We merged the pruned reference data across chromosomes and harmonized variant IDs with our dataset to identify common variants with 1000 Genomes Project reference panel (205.409 variants). PLINK was used to generate a merged dataset and compute principal components, which were used as covariates in downstream analyses. To generate PCA plots, we read eigenvectors and eigenvalues from PLINK output in R and calculated the proportion and cumulative variance explained by each principal component (PC). We annotated samples by merging with 1000 Genomes population metadata and labeled study samples accordingly. All eQTL cohort samples clustered with

European populations, primarily UK, thus genotype PCs were not included as covariates in downstream analyses (Figure S15). Power analysis for the eQTL analysis was performed using the R package powerEQTL^92^ (*powerEQTL.SLR*) across different MAF, slopes and sample sizes (Figure S1).

### Mapping *cis*-eQTL

For the eQTL analysis, we included WGS data and matched RNA-seq tissue-specific expression data from the same patients (nL=L321 osteoarthritis patients across all tissues). For eQTL analyses the gene expression count matrix was normalized using the trimmed mean of M-values (TMM) normalization^93^ (between samples normalization) and then inverse-normal transformed (across genes normalization). This procedure was performed separately per tissue (cartilage, synovium and fat pad). The resulting matrices were then used to estimate PEER factors^94^ representing hidden data variation.

We tested the association of each PEER factor with known covariates. For categorical covariates analysis of variance (ANOVA) was performed between each PEER factor and each covariate via the aov function in R. From the resulting ANOVA summary, the F-statistic and associated p-value (Pr(>F)) were extracted to assess statistical significance. For numeric covariates, a linear regression model was fitted using the lm function in R with PEER as the dependent variable and the covariate as the independent variable. From the regression summary, the t-statistic and p-value corresponding to the covariate were extracted to determine the strength and significance of the association (Figures S16-19).

The number of PEER factors included per tissue in the eQTL model was determined according to GTEx^35^ recommendations on the sample size (low-grade cartilage: 45, high-grade cartilage: 30, synovium: 45, fat pad: 15) [https://github.com/broadinstitute/gtex-pipeline/tree/master/qtl]. For eQTL analyses we included protein-coding and long non-coding RNA (lncRNA) genes excluding other gene biotypes due to higher quantification uncertainty. We further included genes on the autosomes. The total number of genes per tissue used in eQTL testing was formed as follows: cartilage (both low-grade and high-grade): 16,009, synovium: 14,482, fat pad:15,156. We further defined the transcription start-site (TSS) for each gene based on its most abundant transcript across the tested tissues (transcript abundance quantified using RSEM^95^).

We conducted *cis*-eQTL analysis per tissue using tensorQTL (version 1.0.9)^96^ following the GTEx pipeline [https://github.com/broadinstitute/gtex-pipeline/tree/master/qtl]. In brief, this method uses linear regression to test for associations between variant–gene pairs in a specified window (1-Mb specified for *cis-*eQTL) within the transcription start site of a gene (nominal pass). The nominal p-values were estimated for every variant–gene pair with the following model:

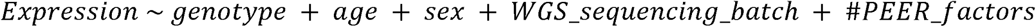

Out of 321 WGS samples, 247 were extracted in the first batch and 74 in the second sequencing batch. The number (#PEER_factors) was determined per tissue.

To test for association between gene expression and the lead variant (top in -*cis*) we used tensorQTL with the implemented permutation scheme specifying 1,000 permutations per gene. This way, we calculated the nominal p value threshold per gene and the gene-level q values corrected for multiple testing (Storey-Tibshirani). To identify eGenes, we performed q value correction of the permutation p values for the lead associated variant per gene at a threshold of 0.05. To identify significant eQTL-gene pairs we filtered the nominal output for the pairs below the gene-specific p value threshold. We further identified conditionally independent eQTL by performing a stepwise regression procedure as described in GTEx^35^.

### Mapping *trans*-eQTL

We performed *trans-*eQTL analysis per tissue using MatrixeQTL^97^. Expression and genotype input data were prepared as described for the *cis-*eQTL analysis. The same covariates and number of PEER factors were included in the final model. We performed *trans-*eQTL analysis on all variants whose distance to the genes’s TSS was more than 5Mb. Given the large number of tests, only gene-SNP pairs with p-value < 10^−6^ were retained. Similar to the *cis*-eQTL analysis, and as previously reported^35^, we calculated gene-level significance as follows:

- For each gene, we selected the most significant *trans*-eQTL (i.e., SNP-gene pair with the lowest nominal p-value) across chromosomes 1–22.
- The nominal p-values were multiplied by 10^6^ to correct for the number of expected independent tests for a genome-wide association analysis at MAF 5% in a European population. Corrected p-values were capped at 1.
- FDR correction on the list of corrected p-values was performed using the qvalue package. *Trans*-eQTLs were defined as those with q-value < 0.05.
- To identify per-gene *trans*-eQTLs, a genome-wide significance threshold was calculated. We identified the largest p-value among significant *trans*-eQTLs (q-value ≤ 0.05) and the smallest p-value among non-significant *trans*-eQTLs (q-value > 0.05). We then took the midpoint between these two values to define a conservative genome-wide p-value threshold for significance at FDR 5%.

Finally, we removed potential false positive *trans*-eQTLs caused by read cross-mapping. To this end, we utilized publicly available cross-mappability resources for the GRCh38 reference genome and excluded all *trans-*eQTL-gene pairs that had a mappability score less than 0.8^98^. We identified conditionally independent *trans*-eQTLs for each analyzed tissue by clumping the significant trans-eQTLs based on LD. We used PLINK^91^ to perform clumping on a window of 1 Mb and a strict LD threshold r^2^ < 0.001.

### *Trans*-eQTL transcription factor binding motif disruption analysis

To assess whether *trans*-eQTLs disrupt transcription factor binding motifs (TFBMs), we used the Bioconductor package motifbreakR^99^ (version 2.20). We filtered the TF motif database to include only human motifs (organism == “Hsapiens”) and restricted it to high-confidence data sources: HOCOMOCOv11-core-A, jaspar2022, JASPAR_CORE, and SwissRegulon. We ran motifbreakR() using the following parameters: filterp = TRUE, method = “ic” (information content), threshold = 1e-5, and a uniform background nucleotide distribution bkg = c(A=0.25, C=0.25, G=0.25, T=0.25). P-values for reference and alternative alleles were computed using calculatePvalue() with granularity = 1e-4. We retained only strong TFBM disruptions by filtering for results with Refpvalue < 0.05, Altpvalue < 0.05, effect == “strong”, and pvalueEffect == “strong”. For each trans-eQTL, we recorded the disrupted TF motif and the associated target eGene to generate candidate TF–eGene regulatory links.

### Comparison with existing *cis-*eQTL maps

To find newly reported associations we compared the significant eQTL-gene pairs with published eQTL studies from Streinberg et al^14^ and Jiang et al.^16^. Both of these studies report eQTL associations on reference genome build 37 while our eQTL maps are on build 38. To establish a fair comparison, we converted the variant positions to the corresponding Reference SNP cluster ID (rsID) as rsIDs do not depend on a reference genome. We mapped variants’ genomic positions to rsIDs using dbSNP database^100^ available through Bioconductor package SNPlocs.Hsapiens.dbSNP144.GRCh37.

We further evaluated the association of effect sizes (slopes) for *cis-*eQTLs by conducting a linear regression analysis of common eQTL effect sizes between our study and that of Steinberg et al.^14^, which was performed in a subset of individuals from our study, all of European ancestry. Specifically, we analyzed 46,433 common *cis-*eQTLs with rsIDs in synovium, 63,294 in low-grade cartilage, and 79,826 in high-grade cartilage. For each tissue type, we fit a linear regression model with the effect sizes from our study as the predictor variable and those from Steinberg et al.^14^ as the outcome variable. From this model, we extracted the R-squared value to quantify the strength of association and the p-value to evaluate statistical significance. To visualize these associations, we used ggplot2^101^ to generate scatter plots with fitted regression lines and confidence intervals for each tissue type.

### Identification of tissue-sharing and single-tissue eQTLs

To assess tissue-specific and shared eQTL patterns, we first explored eQTL effects detection in only one tissue due to tissue-specific gene expression (i.e., eQTLs in genes not common across tissues). To formally assess the presence of unique eQTL effects for genes that are commonly expressed in all disease tissues we applied multivariate adaptive shrinkage using MashR^102^ on the top *cis*-eQTLs for each gene across four tissues, focusing on genes expressed in all tissues (N = 13,181). We input the estimated effect sizes and their associated standard errors, along with 59,310 randomly selected SNP-gene pairs tested across all tissues, to fit the Mash model as previously done ( https://github.com/stephenslab/gtex-eqtls). MashR’s output of effect size estimates and local false sign rates (LFSR), were used as measures of *cis*-eQTL magnitude and significance, respectively. We set an LFSR threshold of < 0.05 to identify significant eQTLs within each tissue. Shared *cis*-eQTLs were defined as those with an LFSR < 0.05 and an effect size within a two-fold range of the strongest effect observed across tissues. To identify tissue-specific *cis*-eQTLs, we implemented a filtering procedure that required: (1) LFSR < 0.05 in a single disease tissue; (2) LFSR ≥ 0.1 in all other tissues; (3) a ≥ two-fold larger absolute effect size in the single tissue compared to all others; and (4) a minimum absolute effect size of 0.1. This ensured eQTLs with unique effect in one tissue.

### Variant annotation of eQTL

We annotated all variants tested in eQTL analyses for functional impact on known genes using the ANNOVAR tool (https://annovar.openbioinformatics.org/en/latest/)^103^. Regulatory annotations were drawn from the Ensembl Regulatory Build^104^, covering features such as enhancers, promoters, promoter-flanking regions, transcription factor binding sites, CTCF binding sites, and open chromatin regions. For low- and high-grade cartilage tissue, we further utilized enhancer-promoter loop coordinates specific to osteoarthritis cartilage, as mapped by chromatin accessibility Hi-C data from Bittner et al.^54^. From this Hi-C map (hg38 build), we extracted genomic coordinates for: (1) chondrocyte-specific CTCF binding sites, marking the most active regions within topologically associated domains (TADs); and (2) active chondrocyte promoters and enhancers within Hi-C loops. Active loops were identified using ATAC-seq open chromatin data combined with ChIP-seq peaks for H3K4me3 and H3K27ac signals^54^. We evaluated the distribution of eQTL annotations among both *cis-* and *trans*-eQTLs (Figure S20).

### Genetic colocalization of *cis*-eQTL with GWAS signals

We conducted regional genetic colocalization analyses between eQTL from osteoarthritis primary tissues (low-grade and high-grade cartilage, synovium, and fat pad) and three knee-related osteoarthritis GWAS (knee osteoarthritis, total knee replacement, and osteoarthritis at any site)^6^. Regions to run genetic colocalization were defined as 1 Mb windows on either side of the phenotype-specific osteoarthritis GWAS risk signals defined in Hatzikotoulas et al. ^6^.Colocalization analysis was run in the GWAS risk regions that had at least one study-wise significant eQTL. To perform the analyses, we have used the *coloc.abf* function of the *coloc* R package (version 5.2.2). To overcome possible LD challenges due to for instance unmatched population structure, evidence of colocalization was set at a posterior probability of a shared causal variant (PP4) >= 0.8 or PP4 > 0.6 and PP4/PP3 > 2, where PP3 denotes the posterior probability of both traits having a distinct causal variant in the genomic region. For each colocalized genomic locus, we calculated a 95% credible set for the shared causal variant by taking the cumulative sum of the variants’ posterior probabilities to be causal conditional on H4 being true. LD between the single-nucleotide polymorphisms (SNPs) was calculated using plink (v.2.0 alpha) based on the UK biobank and was used for visualizing the results in regional association plots^105^.

In regions where no evidence of colocalization but slight evidence of shared causal association between both traits (PP4>0.25) was found, we conducted additional analyses using the *coloc.susie* function which relaxes the assumption of a single shared causal variant by fine-mapping each trait and defining multiple credible sets to run colocalization. For this additional analysis, an LD reference panel is required for each investigated trait. For the eQTL dataset, we used the in-sample LD map and for the osteoarthritis GWAS, we used the UK Biobank genotype files to derive an LD reference map^105^. We defined evidence of colocalization as for colo.abf(): PP4 >= 0.8 or PP4 > 0.6 and PP4/PP3 > 2. Given the lack of credible set information available in coloc.susie, we defined the lead signals from the GWAS and the eQTL as candidates for the lead colocalizing variant.

### Overlap between variants in the 95% credible set of the colocalization and HiC data from chondrocytes

We defined candidates for the shared causal variants as the 95% credible set variants from coloc.abf() results as well as all lead signals colocalizing in coloc.susie(). For each locus showing evidence of colocalization between cis-eQTL data and knee-related osteoarthritis, we mapped all the candidates for the shared causal variants to active enhancer-promoter loops from primary knee osteoarthritis chondrocytes^54^. To identify the target genes of the active promoters, the authors overlaid the extended promoter region with gene regions from ENSEMBL^106^.

### Mendelian randomization analyses between *cis*-eQTL and knee-related osteoarthritis

To evaluate putative causal effects of gene expression levels of colocalizing genes on knee-related osteoarthritis, we conducted Mendelian randomization analyses^58^. As exposures, we used the generated *cis*-eQTL data from the colocalizing genes derived from osteoarthritis primary tissues (low-grade and high-grade cartilage, synovium, and fat pad), and as the outcome, we employed GWAS summary statistics from knee osteoarthritis, osteoarthritis at any site, and total knee replacement^6^. Instrumental variables for the analyses were identified for each gene and tissue combination from the exposure summary statistics as independent variants significantly associated with gene expression. We defined independence through clumping of all significantly associated variants based on LD using the European reference LD panel from the 1000 Genomes project^89^ within a 10Mb window on either side of the index variants, and with a strict threshold of R^2^=0.001. In cases where an instrumental variable was not present in the outcome data, we conducted an LD-based proxy search using R^2^ > 0.7 in the same LD reference panel as in the clumping step. For the proxy search, we used the *LDlinkR* R package (version 1.3.0). We used the *TwoSampleMR* R package (version 0.5.7) curated by MR-Base^107^ to run the causal inference analysis based on the inverse variance weighted method (IVW) and the Wald ratio if only one instrumental variable was available.

Mendelian randomization relies on certain assumptions, which can partially be validated. Firstly, we ensured that the selected instrumental variables were strongly associated with the exposure by filtering out variants with an F-statistic < 10. The F-statistic was calculated as the ratio between the squared effect size estimates and its squared standard error (beta^2^/se^2^)^58^. In cases where multiple instrumental variables were available, we reanalyzed using the inverse variance weighted method with Steiger-filtered exposure data to assess reverse causation by comparing the direction of causal effect estimates. Additionally, if more than two instrumental variables were available, weighted median and MR-Egger analyses were conducted to assess the direction of effect concordance among different estimates of causal effects. To indicate a lack of evidence of pleiotropy, we required the MR-Egger intercept to be equal to zero (FDR adjusted p-value > 0.05). To assess heterogeneity, we examined the FDR adjusted p-value of the Cochran’s Q-statistic, requiring it to be larger than 0.05 for no evidence of heterogeneity. Finally, we have adjusted the p-values of the causal estimates for multiple testing burdens using FDR correction. Hence, evidence for a causal effect was defined based on the following criteria:

1. FDR adjusted p-value of the Wald ratio or IVW method < 0.05
2. If applicable, concordant direction of effect across different tested methods
3. Lack of evidence of pleiotropy (FDR adjusted MR-Egger intercept p-value > 0.05)
4. Lack of evidence of heterogeneity (FDR adjusted Q-statistic p-value > 0.05)

### Pathway enrichment analysis

We have conducted a pathway-based over-representation analysis for all tissue-specific and shared eGenes using all unique gene among tissues as a background (13,181 shared genes). All analyses were performed using ConsensusPathDB (http://cpdb.molgen.mpg.de/)^53^ for both pathways and Gene Ontology terms using the biological process until level five. We further performed enrichment analysis for the genes showing evidence of colocalization with knee-related osteoarthritis phenotypes. For this analysis, we used as the background 9,758 genes that had an eQTL in at least one of the four analyzed osteoarthritis primary tissues. We selected pathways from the biological processes terms of Gene Ontology up to level 5. We required a minimum overlap of two genes for enrichment. The significance threshold was set at false discovery rate (FDR) < 0.05.

### Lookup of prioritized colocalizing genes

To gain further insights into the biology and function of the prioritized genes, we searched these genes in knockout mice and rare and syndromic human diseases databases. Firstly, to look at rare and syndromic diseases linked to the prioritized genes, we extracted data from the Online Mendelian Inheritance in Man (OMIM^61^) (https://omim.org/) via its API. We defined association with musculoskeletal phenotypes if the disease showed any skeletal or muscle soft tissue phenotype. Secondly, we used the batch query functionality of the Mouse Genome Informatics (MGI^60^) database (http://www.informatics.jax.org/) to retrieve knockout mice phenotypes for all prioritized genes. We then screened the output for the following musculoskeletal phenotypes: bone, muscle, skeleton, osteo, arthritis, muscular, joint, body size, growth, skeletal, stature, height, limb, appendage, cartilage, chondrocyte, body length, brachypodia, brachydactyly, brachyphalangia, femur, tibia, ulna, fibula, humerus, radial, spine curvature, posture, vertebral arch and syndactyly.

### Drug targets analysis

We have queried the druggability of the colocalizing genes using the OpenTargets^71^ database (version 23.09), which includes information about 61,264 unique genes. Open Targets defines drugs as any bioactive molecule with drug-like properties, based on the EMBL-EBI ChEMBL database^108^. This definition excludes vaccines, blood products, cell therapies and multi-ingredient drugs. We considered drugs with information on the indication that had completed at least a Phase 2 trial.

### Spatial transcriptomics study participants

The spatial transcriptomics project has been evaluated and approved by the Research Ethics Committee of the University of Tartu (379/M-10, 12th of June 2023). All study participants have voluntarily signed the informed consent form of the study. For spatial transcriptomics analyses three knee osteoarthritis patients from the Orthopaedics Clinic at the Tartu University Hospital were recruited. They were all female with average age 65 (standard deviation 6.78) years and with European ethnicity. They all have been diagnosed with knee osteoarthritis with Kellgren and Lawrence grade 4 (severe) and were hospitalised for the joint replacement surgery due to osteoarthritis. They did not have post-traumatic osteoarthritis , treatment with bisphosphonates, dysplasia, inflammatory arthritis (such as ankylosing spondylitis, rheumatoid arthritis, psoriatic arthritis), gout or lupus.

### Sample collection and preparation

Punch biopsies were taken from the tibial plateau (surgical waste from arthroplasty surgery) immediately after removal from the joint. Biopsies were collected from macroscopically degraded region. All biopsies contained cartilage and subchondral bone tissue layers. Tissues were transferred into 4 % formaldehyde solution at room temperature (RT). Within 2 hours the tissues were cut into smaller pieces (smallest dimension ∼2 mm) and incubated in fresh 4 % formaldehyde solution at RT for at least 72 h with mild agitation. After this, tissues were washed with nuclease free water for 15 min at RT with mild agitation three times and then transferred into 14 % EDTA solution with pH at least 8 for the decalcification. Tissues were incubated in EDTA solution at RT with mild agitation and solution changed daily for altogether 11 days. After that the tissues were washed with nuclease free water for 15 min at RT with mild agitation three times and delivered to Tartu University Hospital Pathology Service laboratory for FFPE block preparation. The FFPE blocks were stored at 4 OC until further use. For conducting the spatial transcriptomics sequencing the tissues were sectioned (5 µm) and placed onto Leica Bond plus slides (Leica Biosystems GmbH, Germany). In parallel the sections were graded based on OARSI grading system and all tissues were at OARSI grade 3.

### Spatial Transcriptomics

Spatial transcriptomics analysis was conducted with Nanostring GeoMx Digital Spatial Profiler platform applying Human Whole Transcriptome Atlas panel (Burker Spatial Biology Inc, USA) and following sequencing on Illumina platform (Illumina Inc, USA). For selecting the areas of illumination (AOIs), the DNA dye signals were used. The AOIs included in the study were selected from the following tissue areas: (1) cartilage superficial zone, (2) cartilage middle zone, (3) cartilage deep zone, (4) calcified cartilage (below tidemark).

### Evaluating the tissue-level specificity of the GO2 eGenes

To evaluate in which osteochondral tissue zones the colocalizing genes are expressed, the following workflow was applied. (1) We looked, which colocalizing genes were detected by Nanostring GeoMX platform in the samples. 97 out of the 117 colocalizing genes were expressed in at least one tissue. (2) We considered the genes not expressed if the probe count was below 6 in at least two samples out of three per zone group^109^. The probe counts can be found in Table S17.

## Data Availability

The full summary statistics of all analyses will be shared through the Musculoskeletal Knowledge Portal (mskkp.org). All software used in this study is available from free repositories or manufacturers as referenced throughout the methods section.

## Acknowledgements

The authors wish to thank Norbert Bittner for providing access to the HiC chondrocyte data and contributing to the discussion for their optimal use. We acknowledge the technical support of Core Facility Genomics at Helmholtz Munich. We thank Dr. Inti Alberto de la Rosa Velasquez, Dr. Peter Lichtner and Dr. Gertrud Eckstein and Dr. Jenny Hankinson for their help in sample handling and additional processing of RNA-seq and WGS data. We also wish to thank Iris Fischer for her helpful contributions during manuscript submission process, and all study participants for their willingness to donate study samples. The work on osteoarthritis cartilage samples was funded by Wellcome Trust (206194).

ER, RM and GLVS have received funding from the European Union’s Horizon 2020 research and innovation programme under Grant Agreement No 101095084 (ENDOTARGET). RM is supported by Estonian Research Council grant PUT (PRG1911) and by Estonian Ministry of Education and Research Funding (TK214). The work on KT and AM is supported by Estonian Research Council grant PSG610.

## Author contributions

Study design: E.Z., J.M.W.; Clinical collection and library preparation: J.M.W., K.M.S., D.S., E.R;, S.S., G.L.V.S., K.T., A.M., R.M., Data Analysis: G.K., A.A., M.T., E.R, P.K.; Interpretation of results: G.K., A.A., E.Z., J.M.W, L.S.; Data visualization: G.K., A.A., M.T; Manuscript drafting: G.K., A.A., E.Z.; Manuscript reviewing and editing: all authors.

## Data and Code availability

The full summary statistics of all analyses will be shared through the Musculoskeletal Knowledge Portal^111^ (mskkp.org). All software used in this study is available from free repositories or manufacturers as referenced throughout the methods section.

