## Supplementary figures and images for "Transcriptome analysis in osteoarthritis primary tissues identifies high-confidence effector genes"

### Supplemental Figure 1

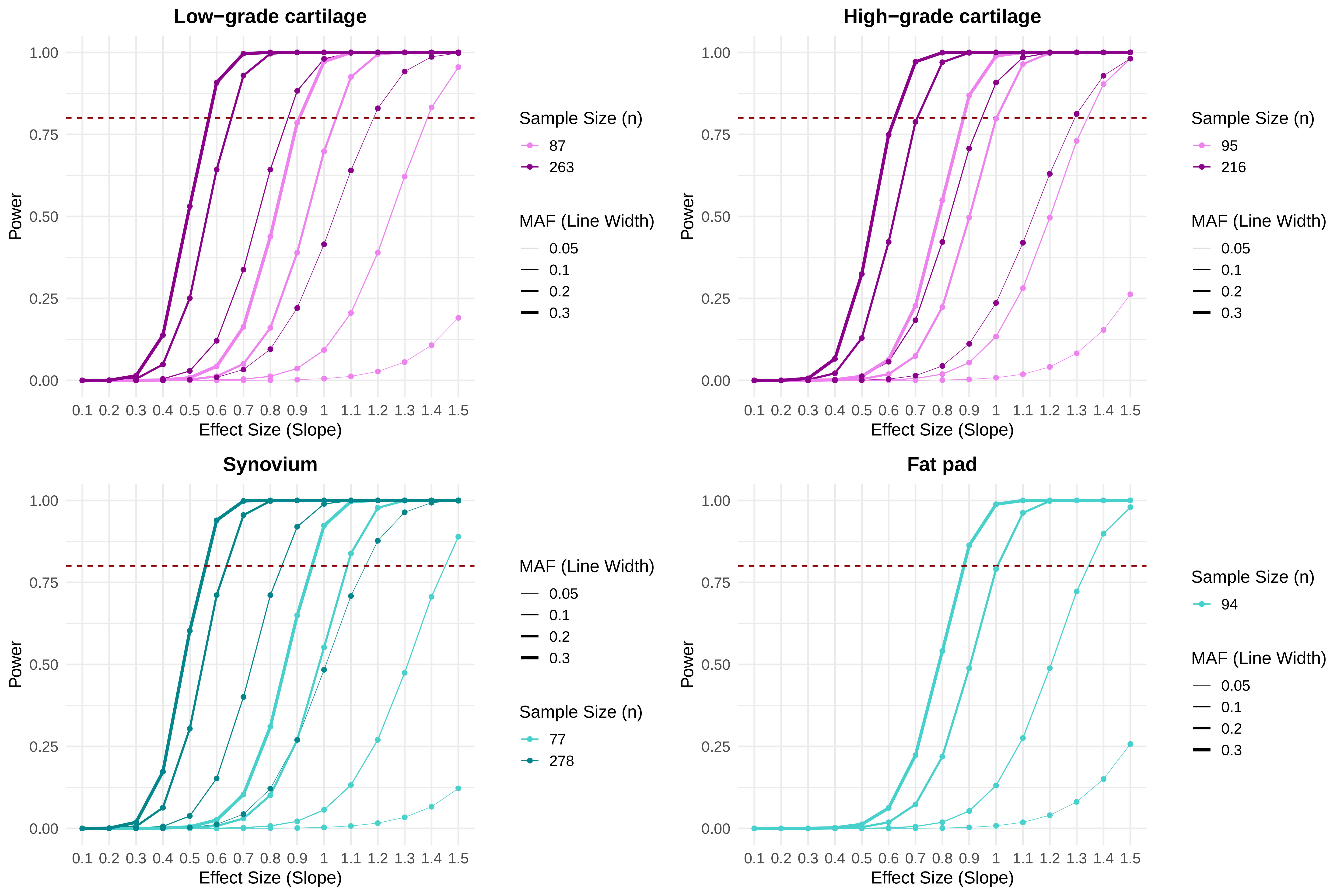

### Supplemental Figure 2

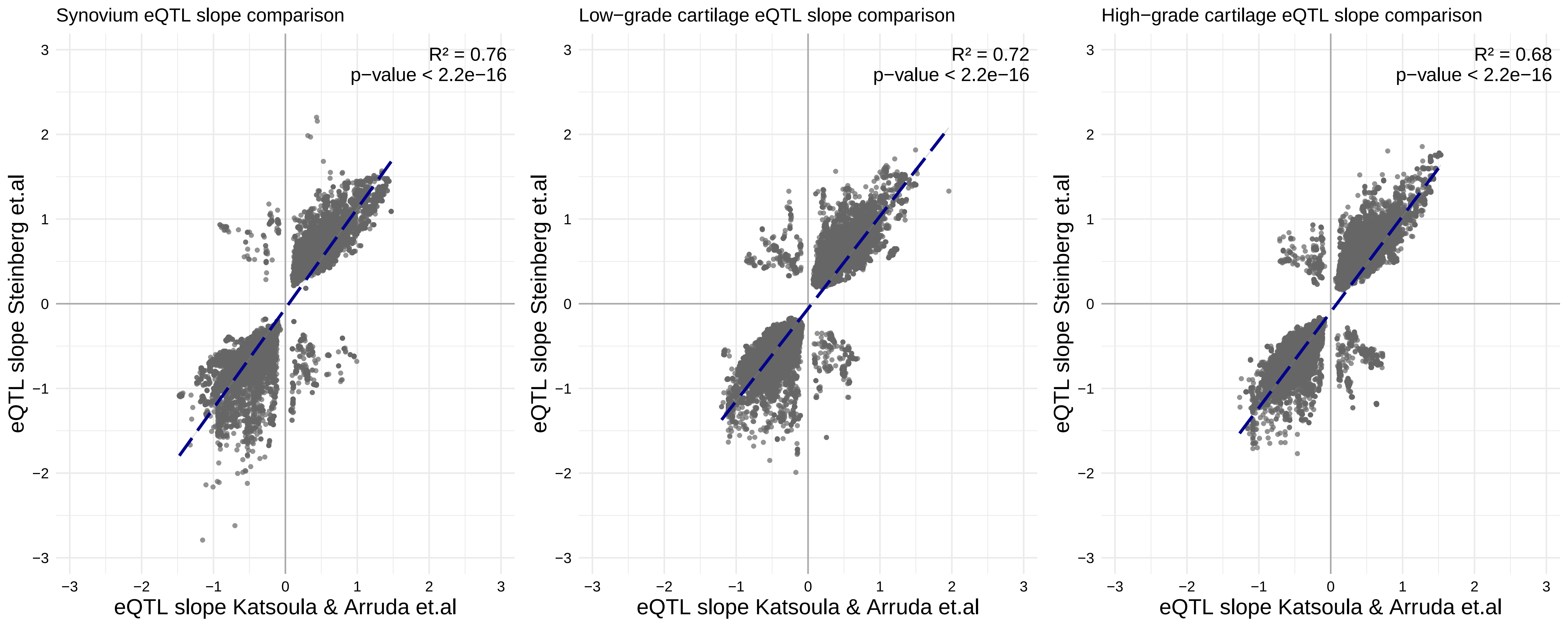

### Supplemental Figure 3

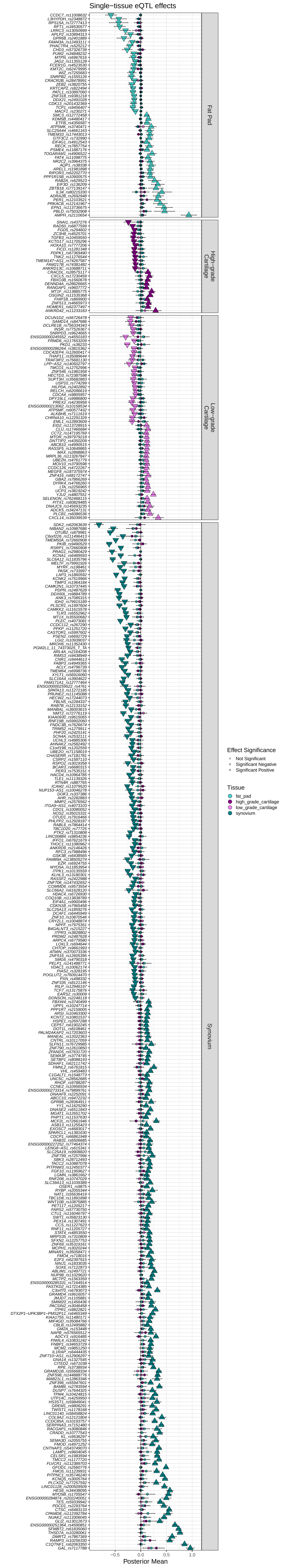

### Supplemental Figure 4

##### Disease−progression eQTLs

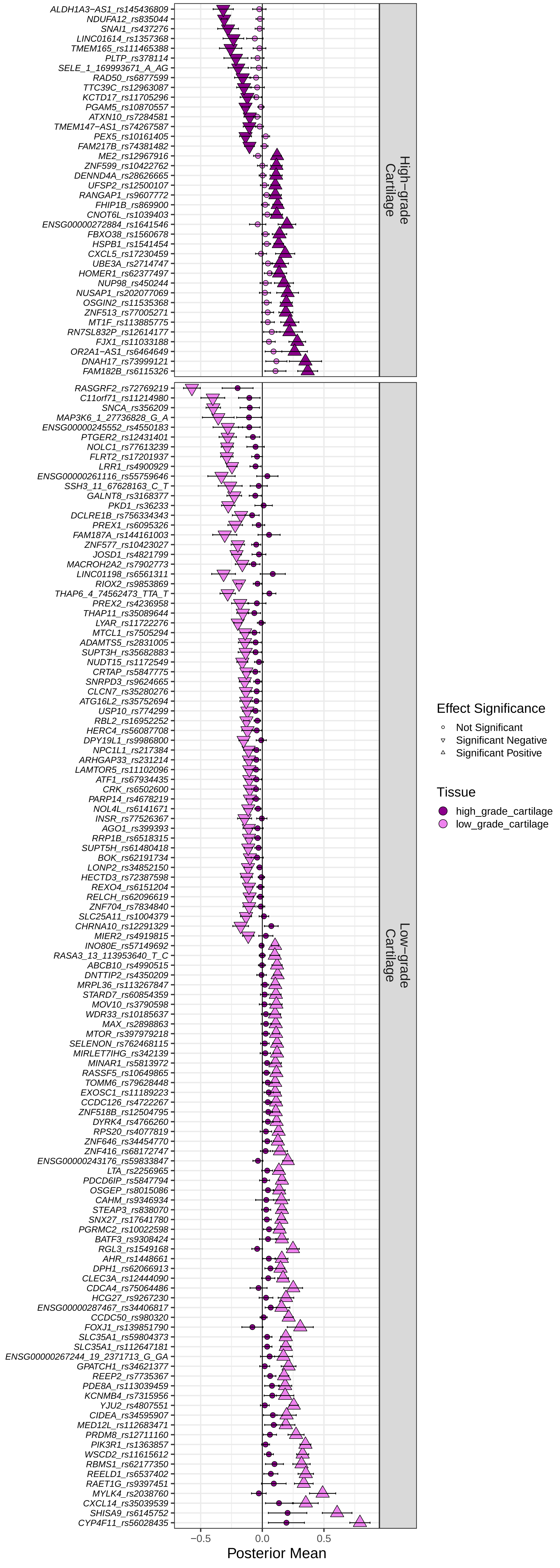

### Supplemental Figure 5

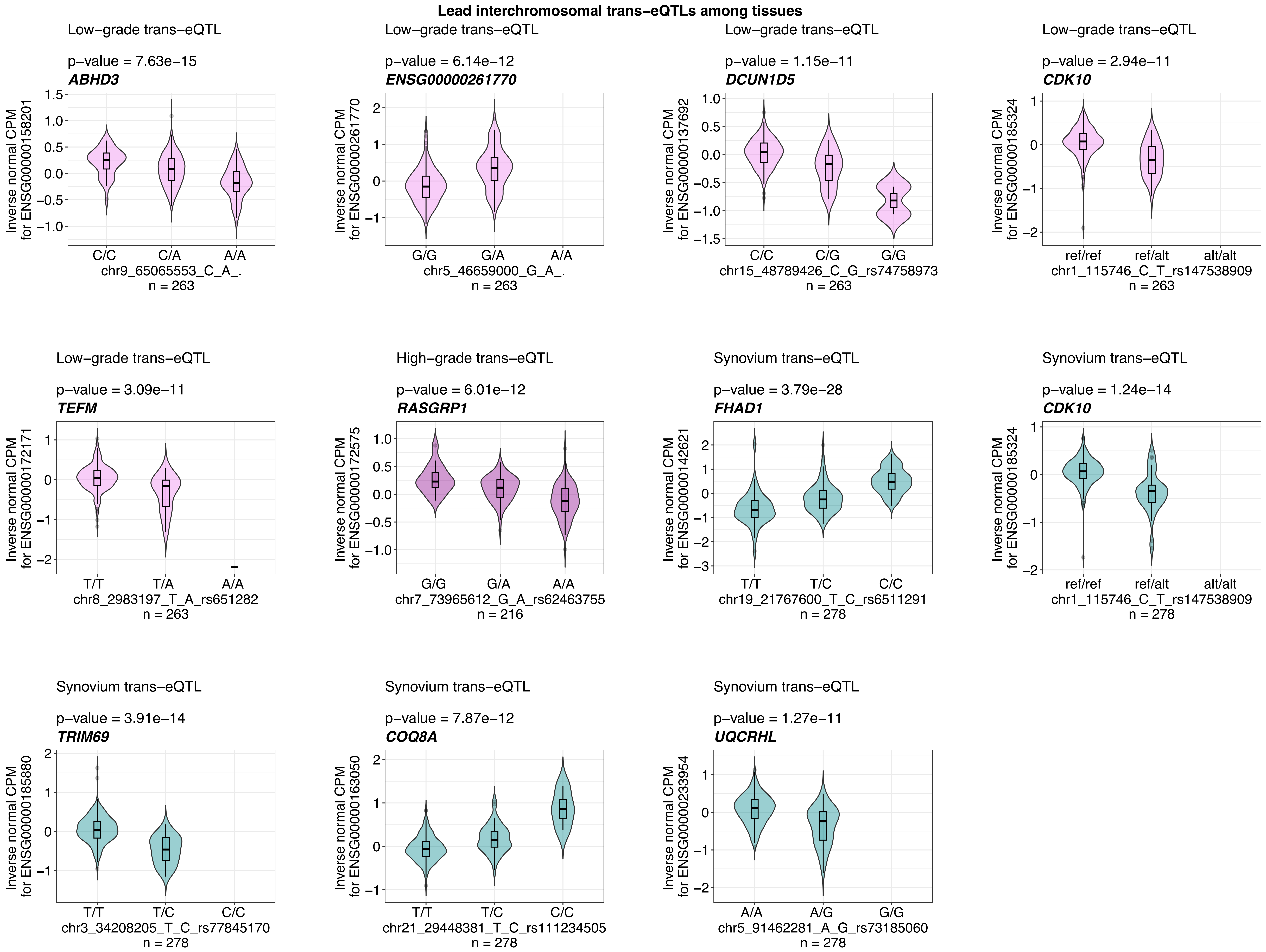

### Supplemental Figure 6

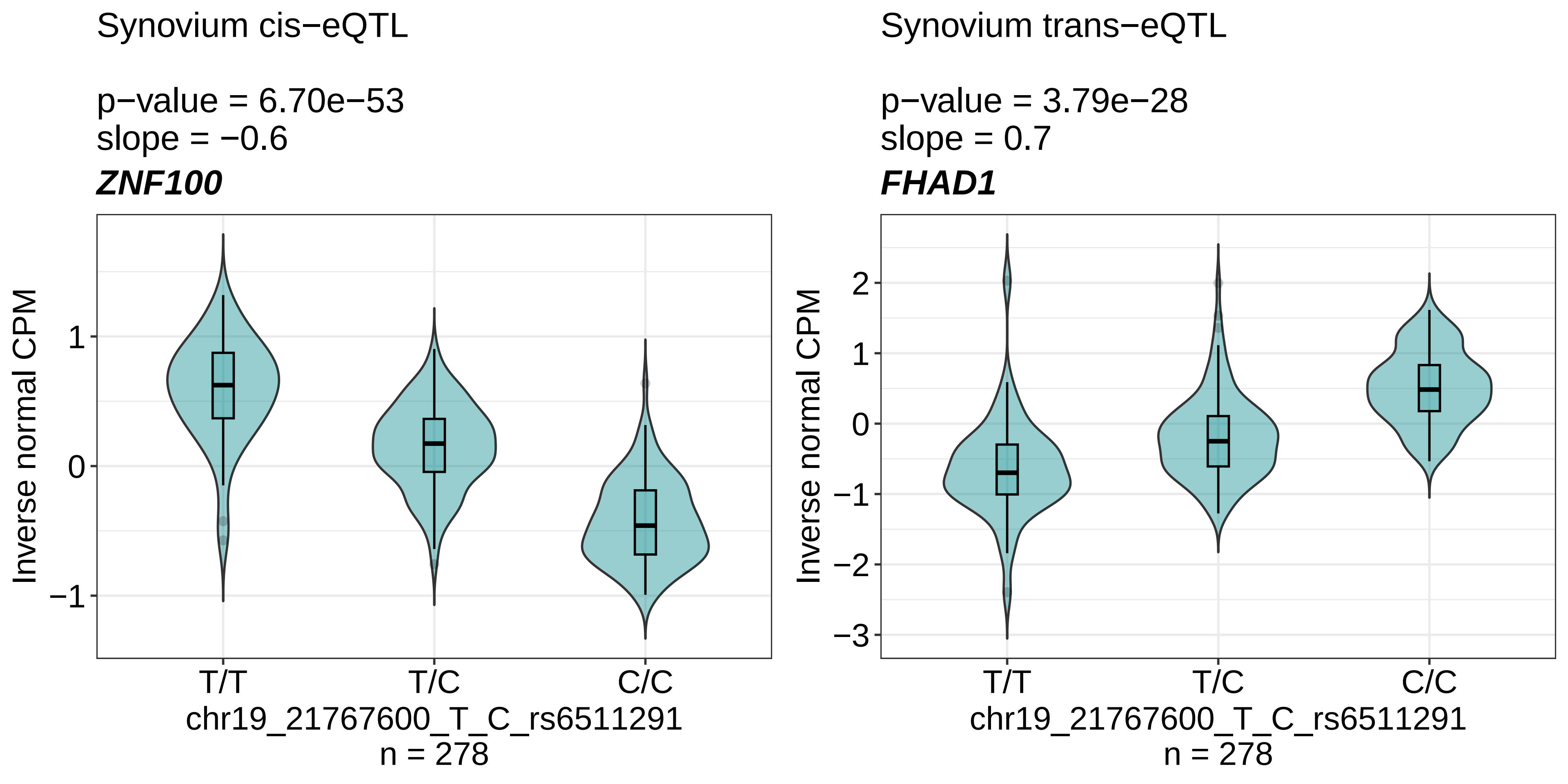

### Supplemental Figure 14

#### PCA plots log CPM normalized data
#### 14,110 genes − 881 samples

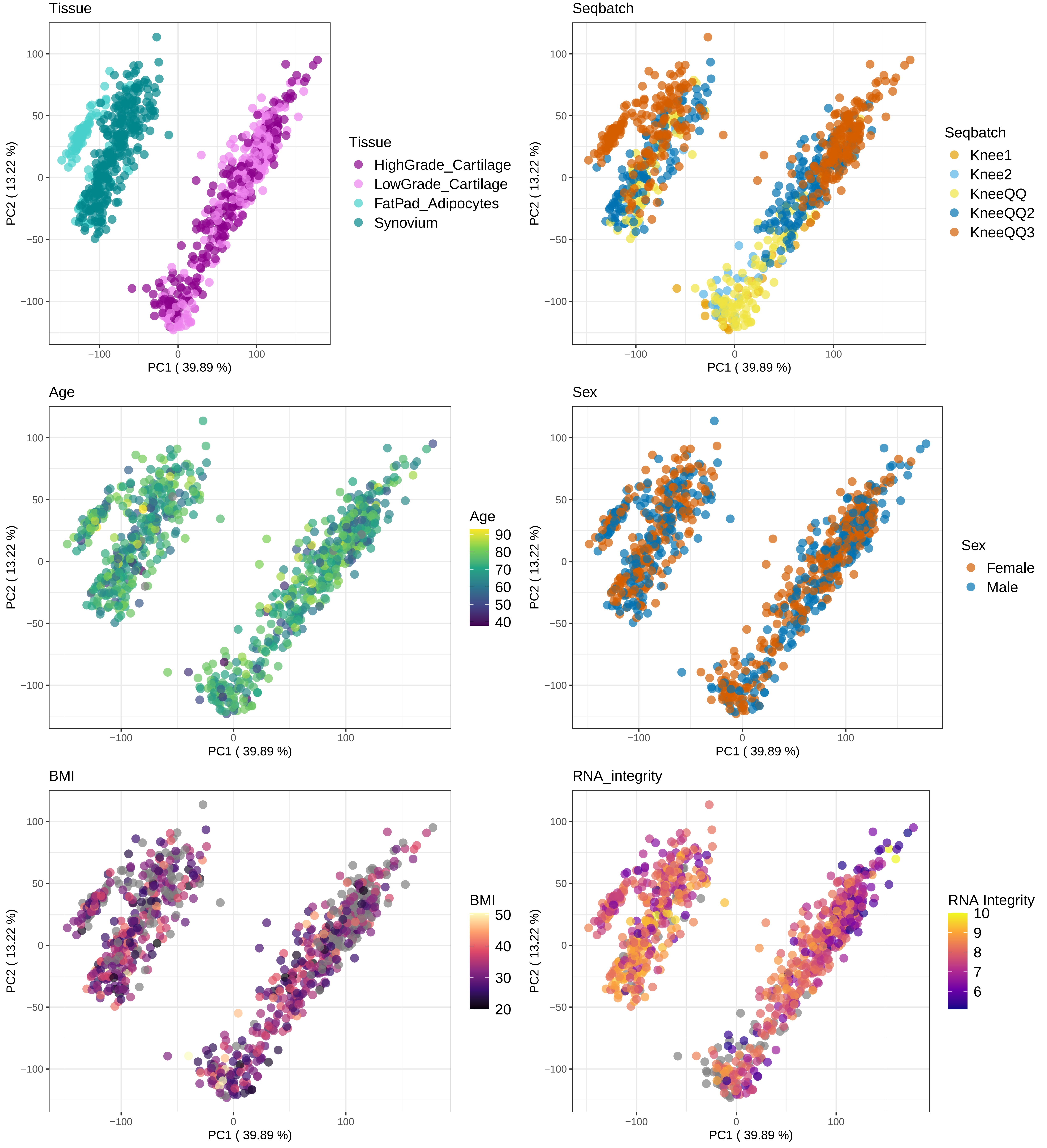

### Supplemental Figure 15

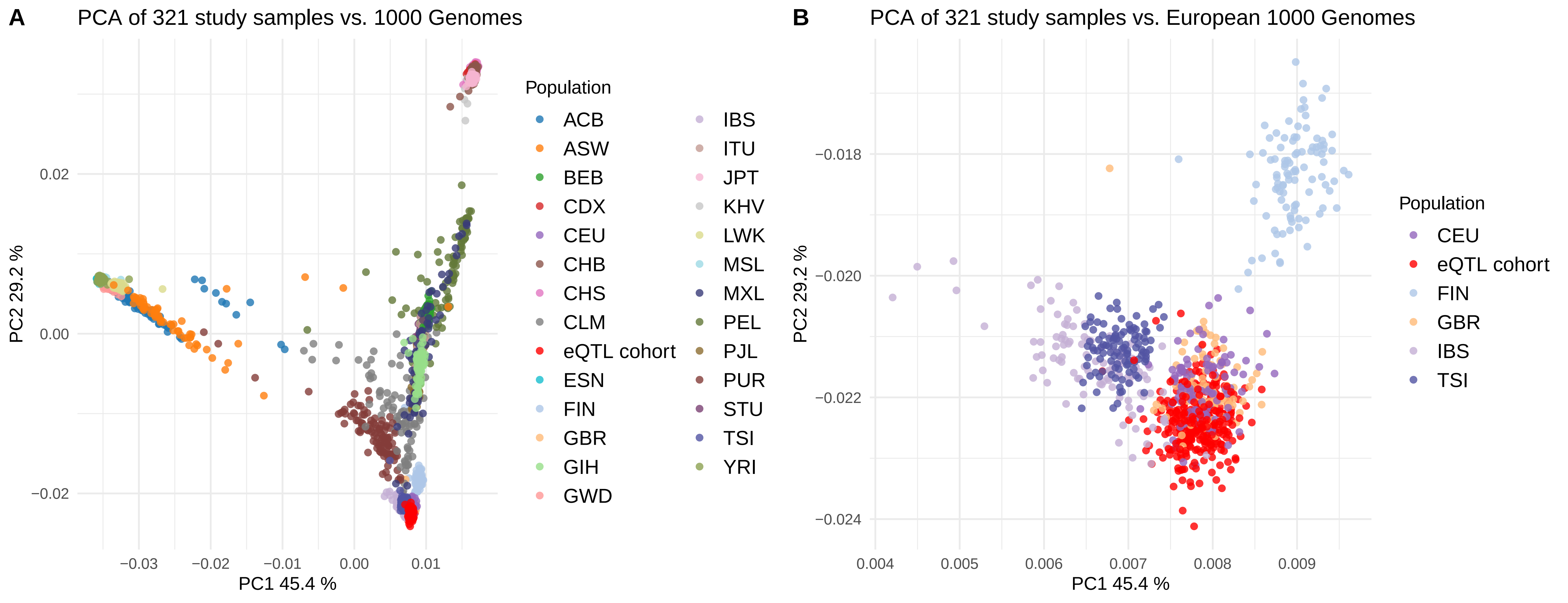

### Supplemental Figure 20

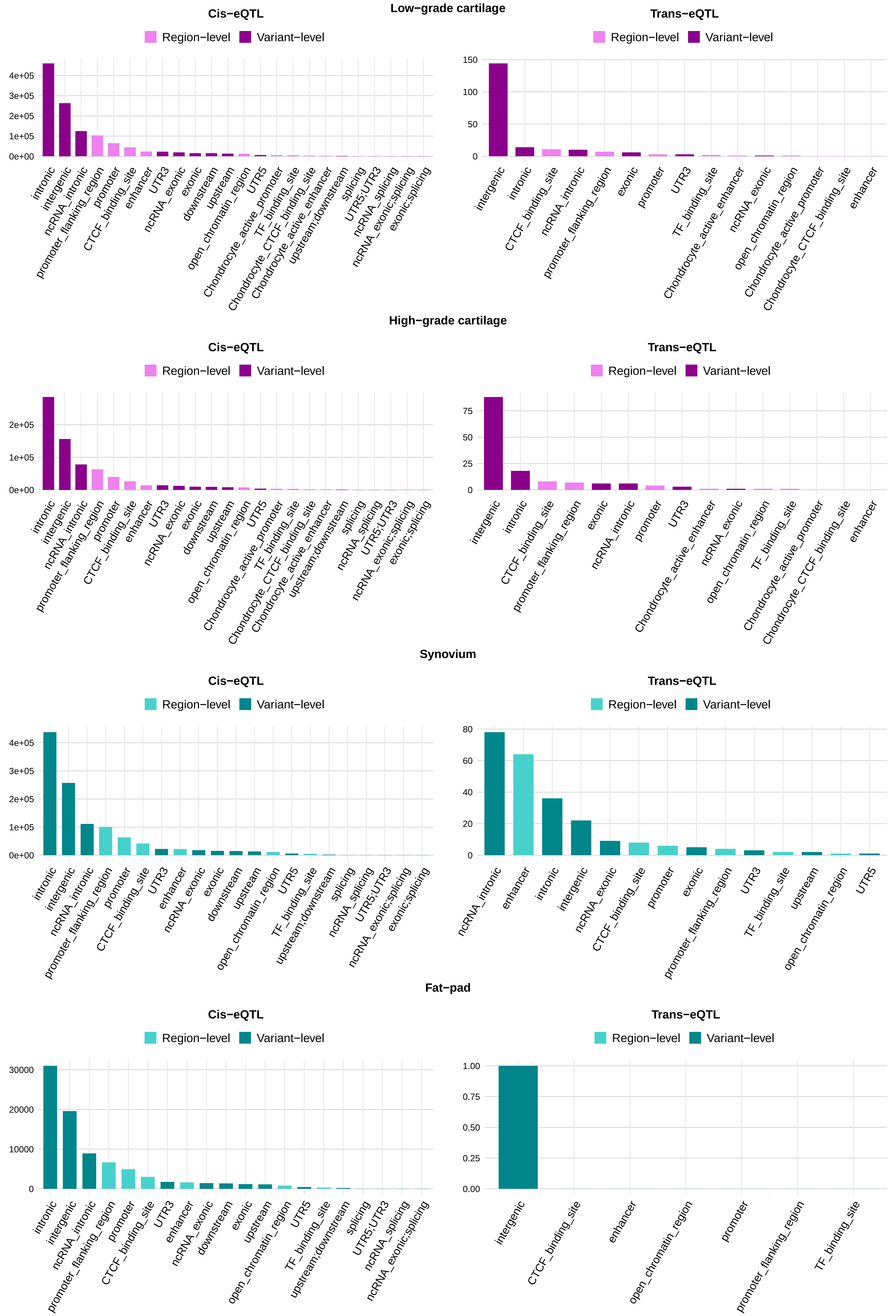

### Supplemental Figures 16-19

#### Figure S16

##### Low-grade cartilage

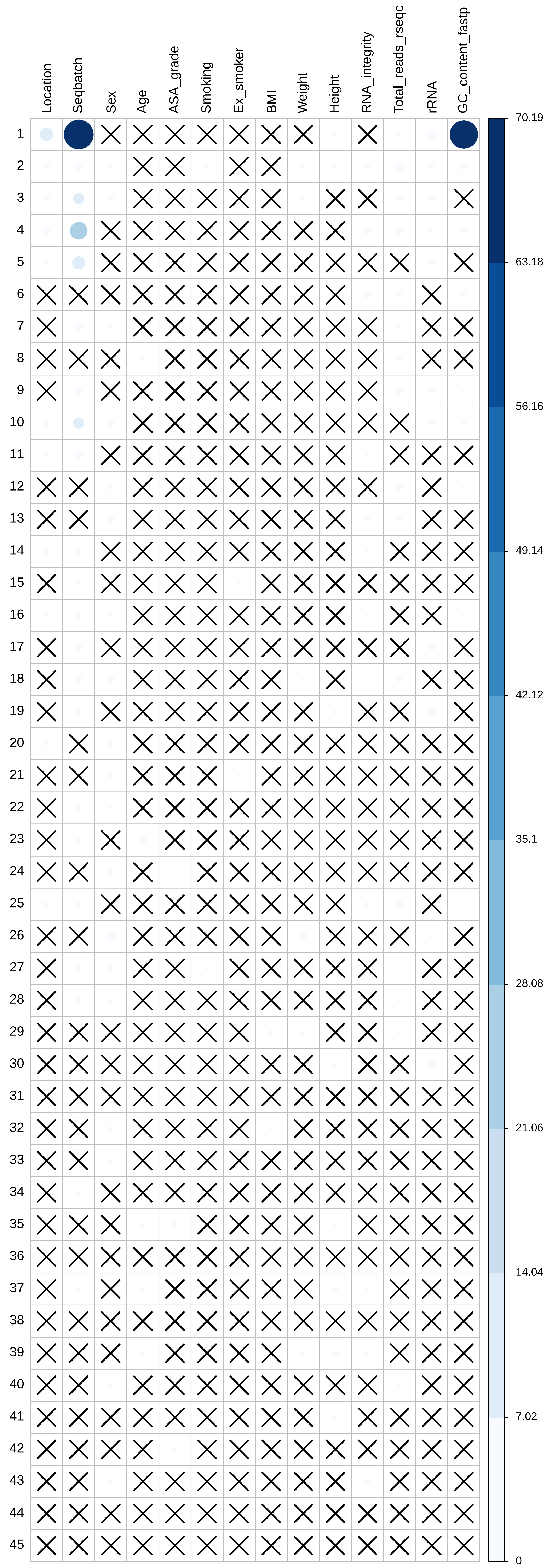

#### Figure S17

High-grade cartilage

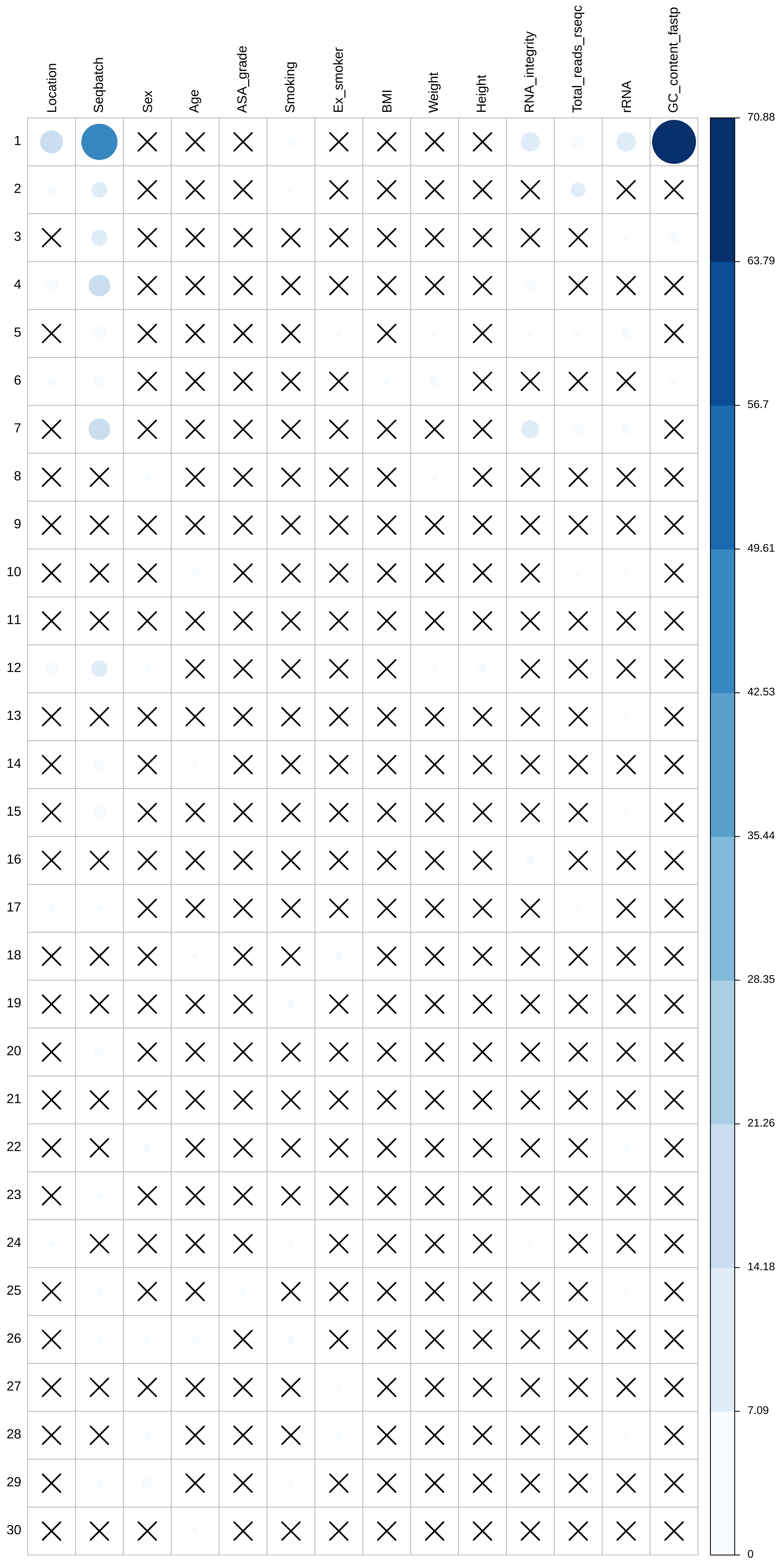

#### Figure S18

Synovium

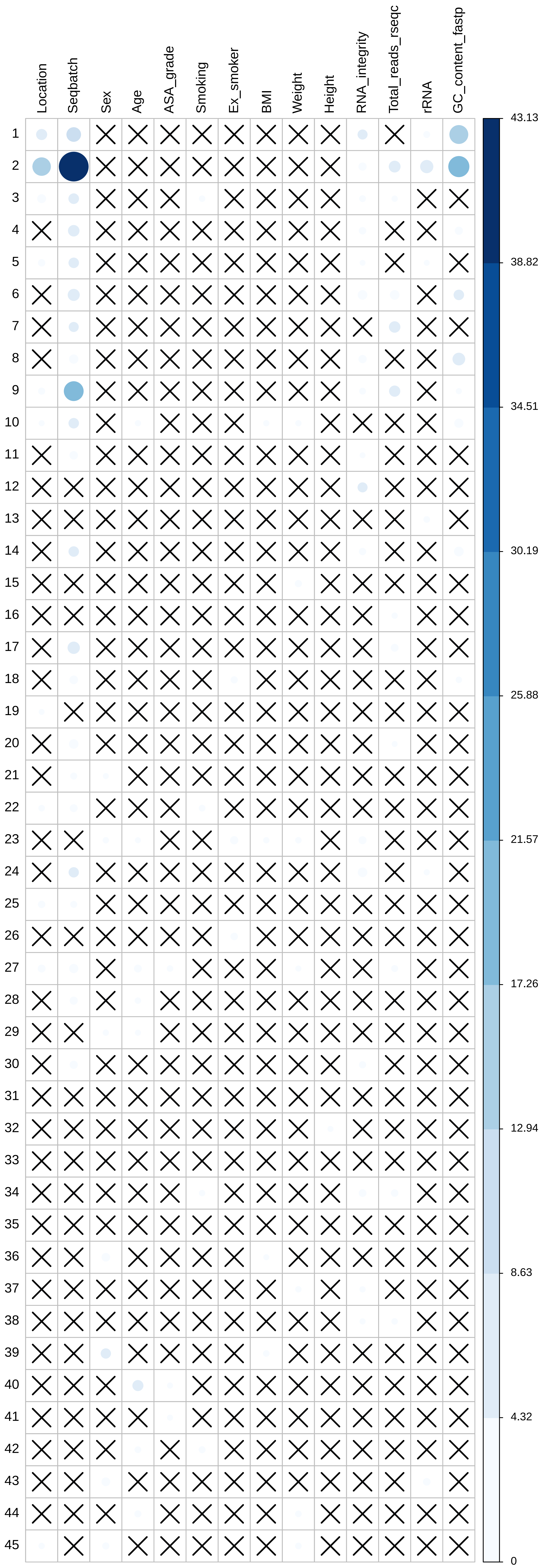

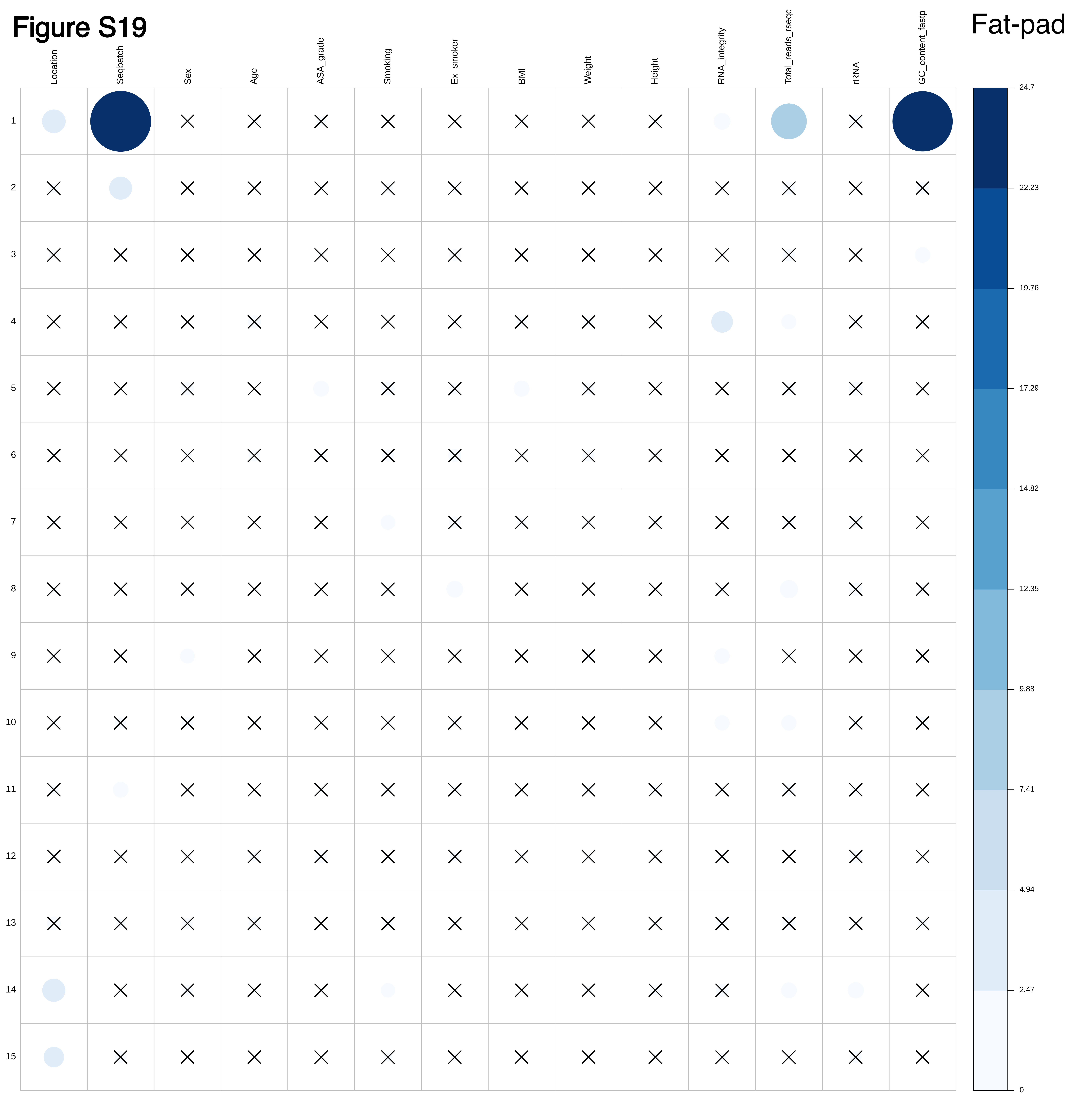
