## Supplemental Figures 7-13 for "Transcriptome analysis in osteoarthritis primary tissues identifies high-confidence effector genes"

**Figures S7-8**: Regional association plots of the genetic loci where a colocalizing eQTL (variant included in the 95% credible set for the shared causal variant from the colocalization analysis with osteoarthritis GWAS) resides within an enhancer of an active promoter-enhancer loop derived from HiC data of primary knee osteoarthritis chondrocytes (*Bittner et al*.) from the same gene (KNEE = knee osteoarthritis; ALLOA = osteoarthritis at any site; TKR = total knee replacement surgery; PP4 = posterior probability of a shared causal variant).

**Figure S7**: *SLC44A2* gene (only the colocalizations with low-grade-cartilage are depicted)
**Figure S8:** *WWP2* gene

#### Figure S7

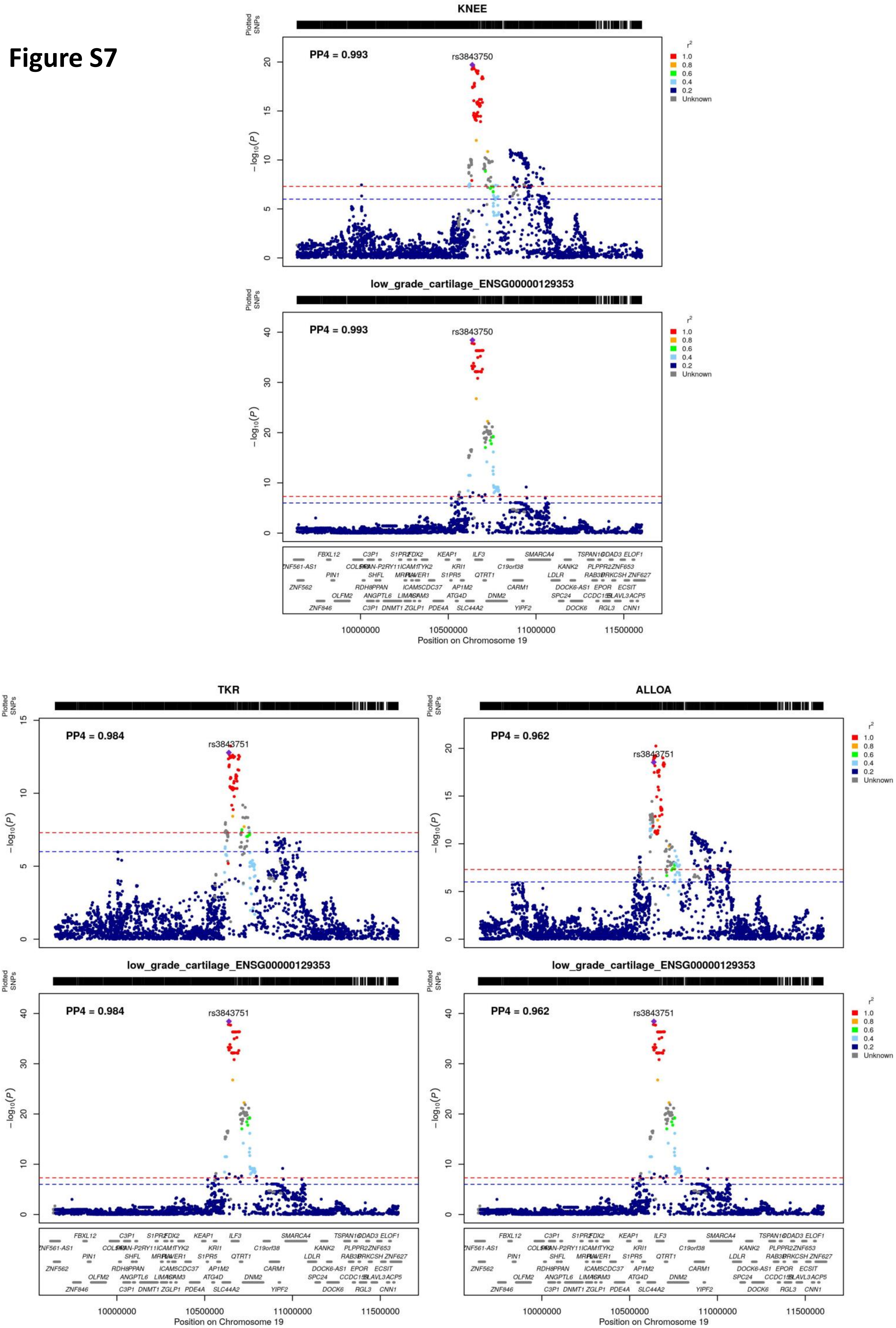
KNEE
PP4 = 0.993
rs3843750
low_grade_cartilage_ENSG00000129353
PP4 = 0.993
rs3843750
Position on Chromosome 19
TKR
PP4 = 0.984
rs3843751
ALLOA
PP4 = 0.962
rs3843751
low_grade_cartilage_ENSG00000129353
PP4 = 0.984
rs3843751
low_grade_cartilage_ENSG00000129353
PP4 = 0.962
rs3843751
Plotted SNPs
$-\log_{10}(P)$
$r^2$
1.0
0.8
0.6
0.4
0.2
Unknown
10000000
10500000
11000000
11500000
Position on Chromosome 19

### Figure S8

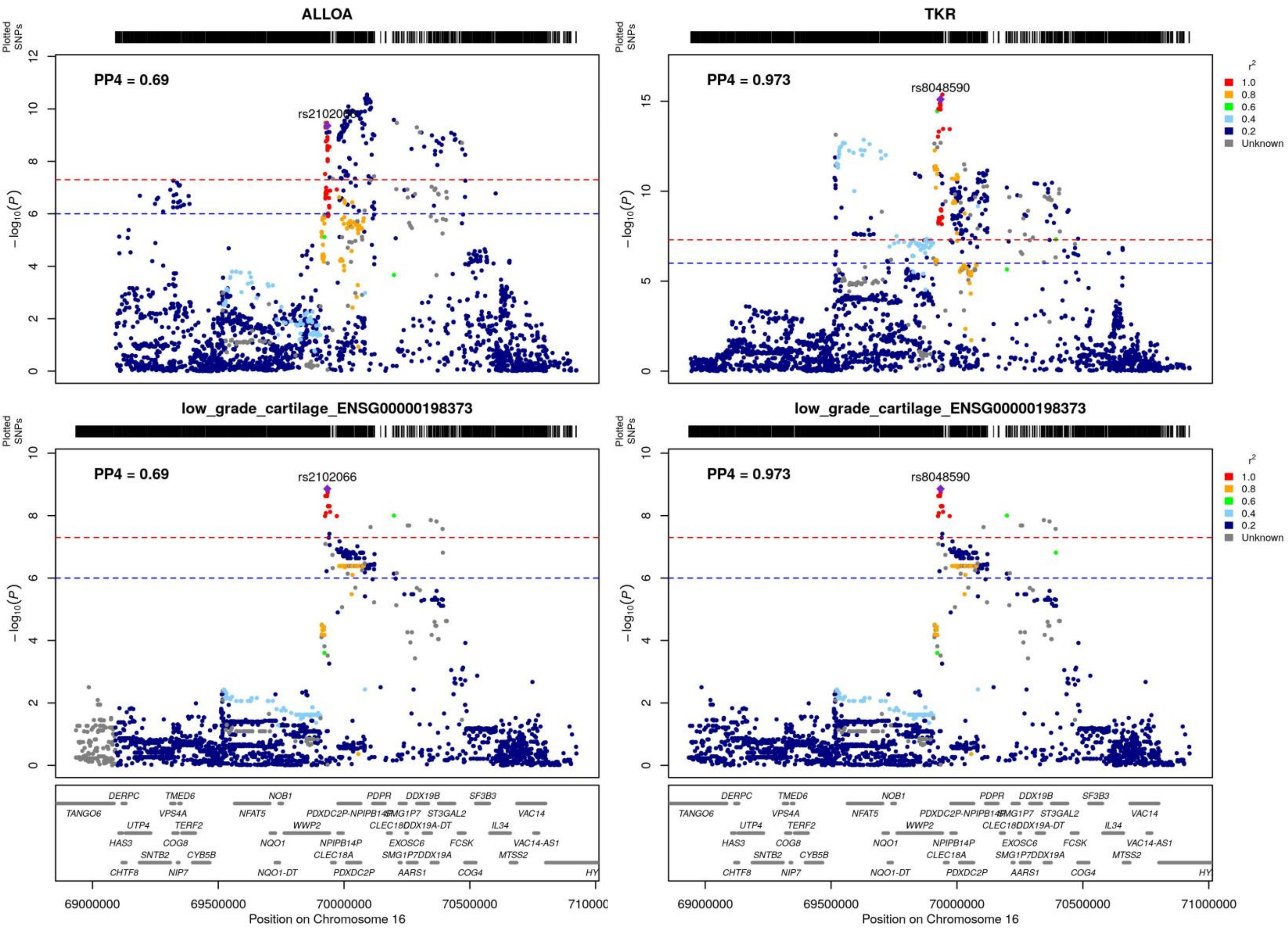
ALLOA
TKR
low_grade_cartilage_ENSG00000198373
low_grade_cartilage_ENSG00000198373
PP4 = 0.69
PP4 = 0.973
rs2102066
rs8048590
Plotted SNPs
$-\log_{10}(P)$
$r^2$
1.0
0.8
0.6
0.4
0.2
Unknown
Position on Chromosome 16
69000000
69500000
70000000
70500000
71000000
DERPC
TMED6
NOB1
PDPR
DDX19B
SF3B3
TANGO6
VPS4A
NFAT5
PDXDC2P-NPIPB14P
SMG1P7
ST3GAL2
VAC14
UTP4
TERF2
WWP2
CLEC18C
DDX19A-DT
IL34
HAS3
COG8
NQO1
NPIPB14P
EXOSC6
FCSK
VAC14-AS1
SNTB2
CYB5B
CLEC18A
SMG1P7
DDX19A
MTSS2
CHTF8
NIP7
NQO1-DT
PDXDC2P
AARS1
COG4

**Figures S9-S13**: Regional association plots of the genetic loci where a colocalizing eQTL (variant included in the 95% credible set for the shared causal variant from the colocalization analysis with osteoarthritis GWAS) resides within a promoter of an active promoter-enhancer loop derived from HiC data of primary knee osteoarthritis chondrocytes (*Bittner et al.*) from the same gene (ALLOA = osteoarthritis at any site; TKR= total knee replacement surgery; KNEE = knee osteoarthritis; PP4 = posterior probability of a shared causal variant).

**Figure S9**: *LGALS3*
**Figure S10**: *WWP2*
**Figure S11**: *FES*
**Figure S12**: *ZNF697*
**Figure S13**: *MSL1*

### Figure S9

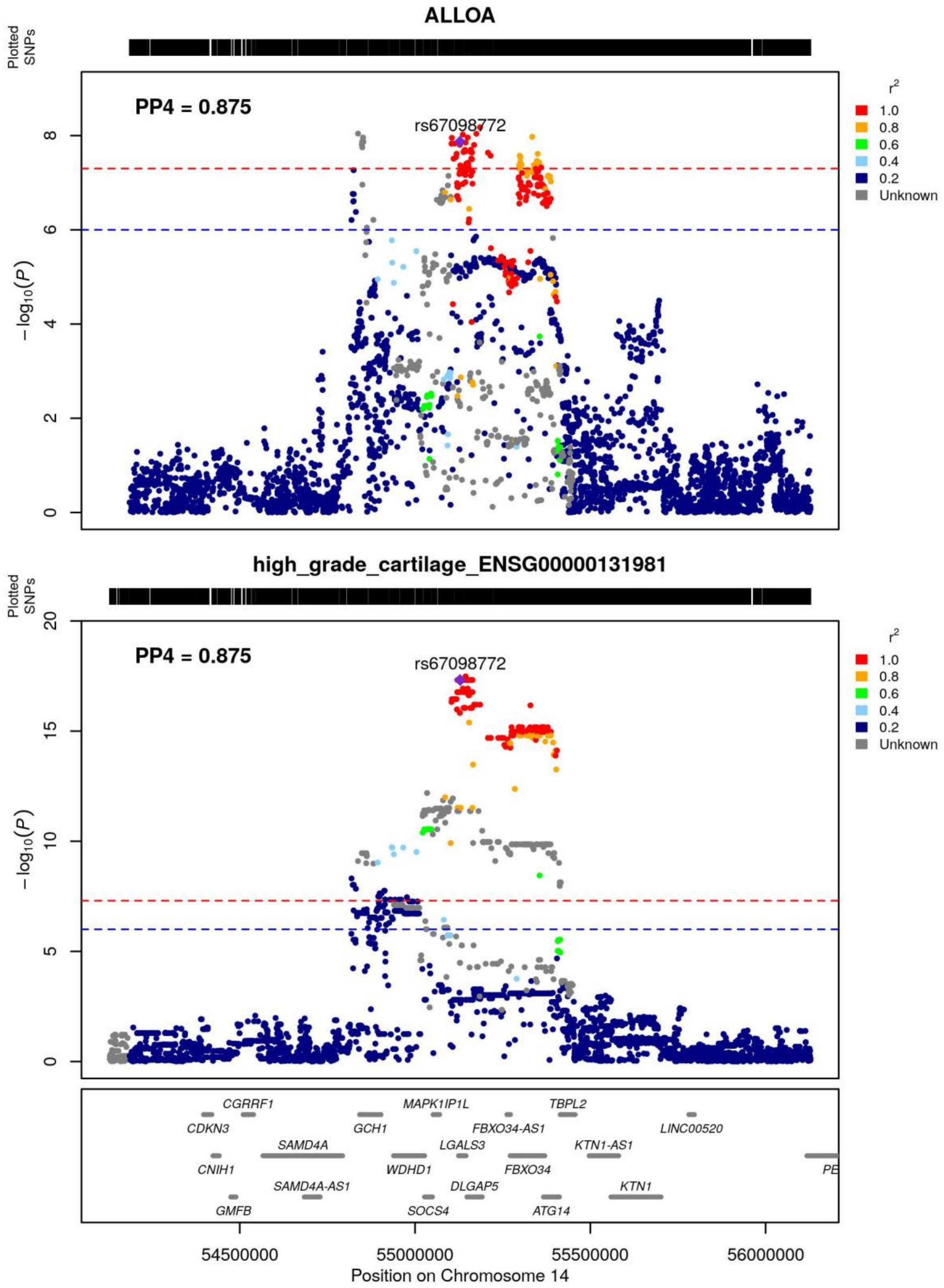
ALLOA
Plotted SNPs
PP4 = 0.875
rs67098772
−log10(P)
r²
1.0
0.8
0.6
0.4
0.2
Unknown
high_grade_cartilage_ENSG00000131981
Plotted SNPs
PP4 = 0.875
rs67098772
−log10(P)
CGRRF1
CDKN3
CNIH1
SAMD4A
SAMD4A-AS1
GMFB
GCH1
MAPK1IP1L
WDHD1
LGALS3
SOCS4
DLGAP5
FBXO34-AS1
FBXO34
TBPL2
ATG14
KTN1-AS1
KTN1
LINC00520
54500000
55000000
55500000
56000000
Position on Chromosome 14

**Figure S10**

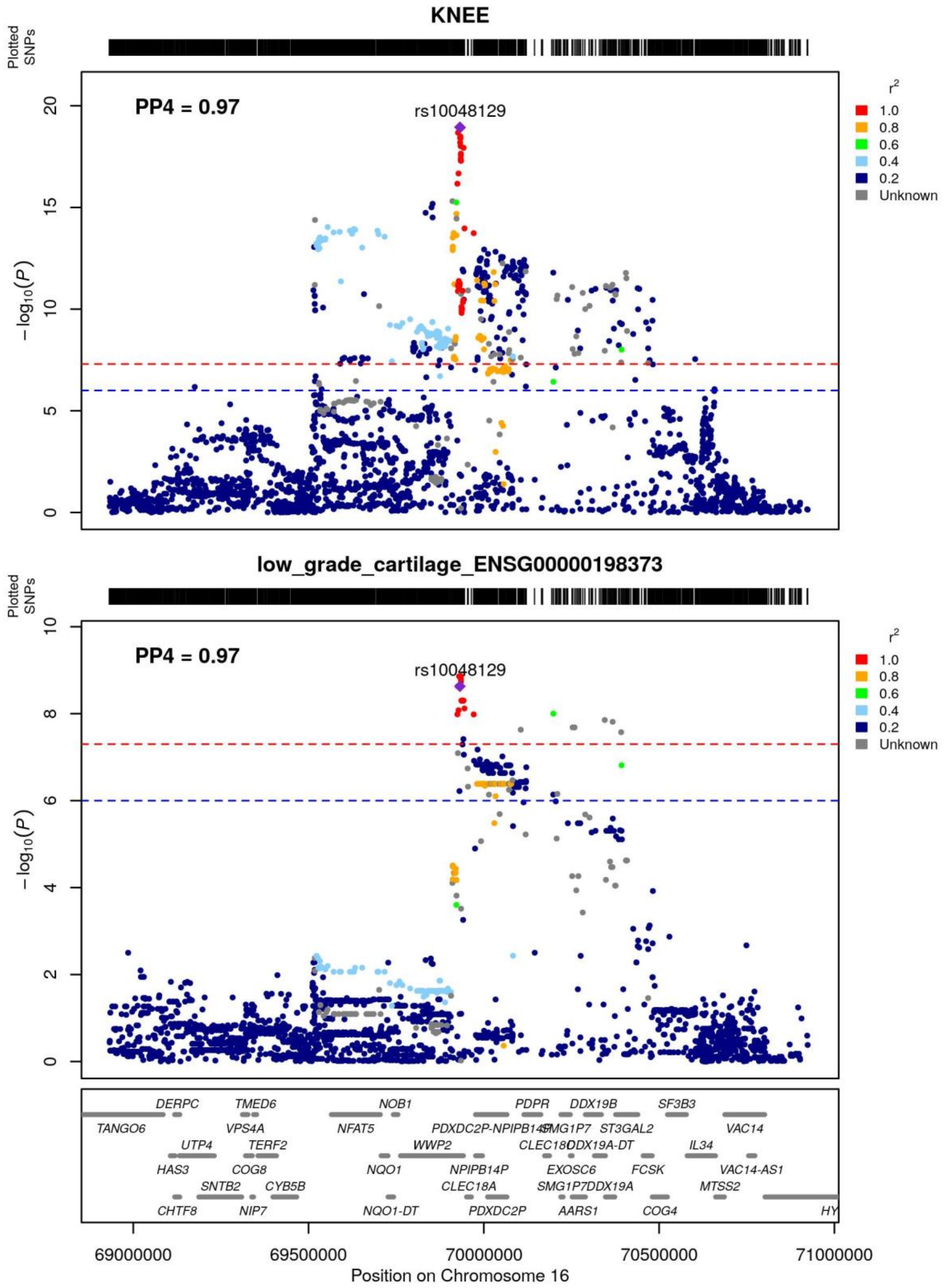

KNEE
Plotted SNPs
PP4 = 0.97
rs10048129
−log10(P)
r2
1.0
0.8
0.6
0.4
0.2
Unknown
low_grade_cartilage_ENSG00000198373
Plotted SNPs
PP4 = 0.97
rs10048129
−log10(P)
DERPC
TMED6
NOB1
PDPR
DDX19B
SF3B3
TANGO6
VPS4A
NFAT5
PDXDC2P-NPIPB14P
SMG1P7
ST3GAL2
VAC14
UTP4
TERF2
WWP2
CLEC18C
DDX19A-DT
IL34
HAS3
COG8
NQO1
NPIPB14P
EXOSC6
FCSK
VAC14-AS1
SNTB2
CYB5B
CLEC18A
SMG1P7
DDX19A
MTSS2
CHTF8
NIP7
NQO1-DT
PDXDC2P
AARS1
COG4
HY
69000000
69500000
70000000
70500000
71000000
Position on Chromosome 16

### Figure S11

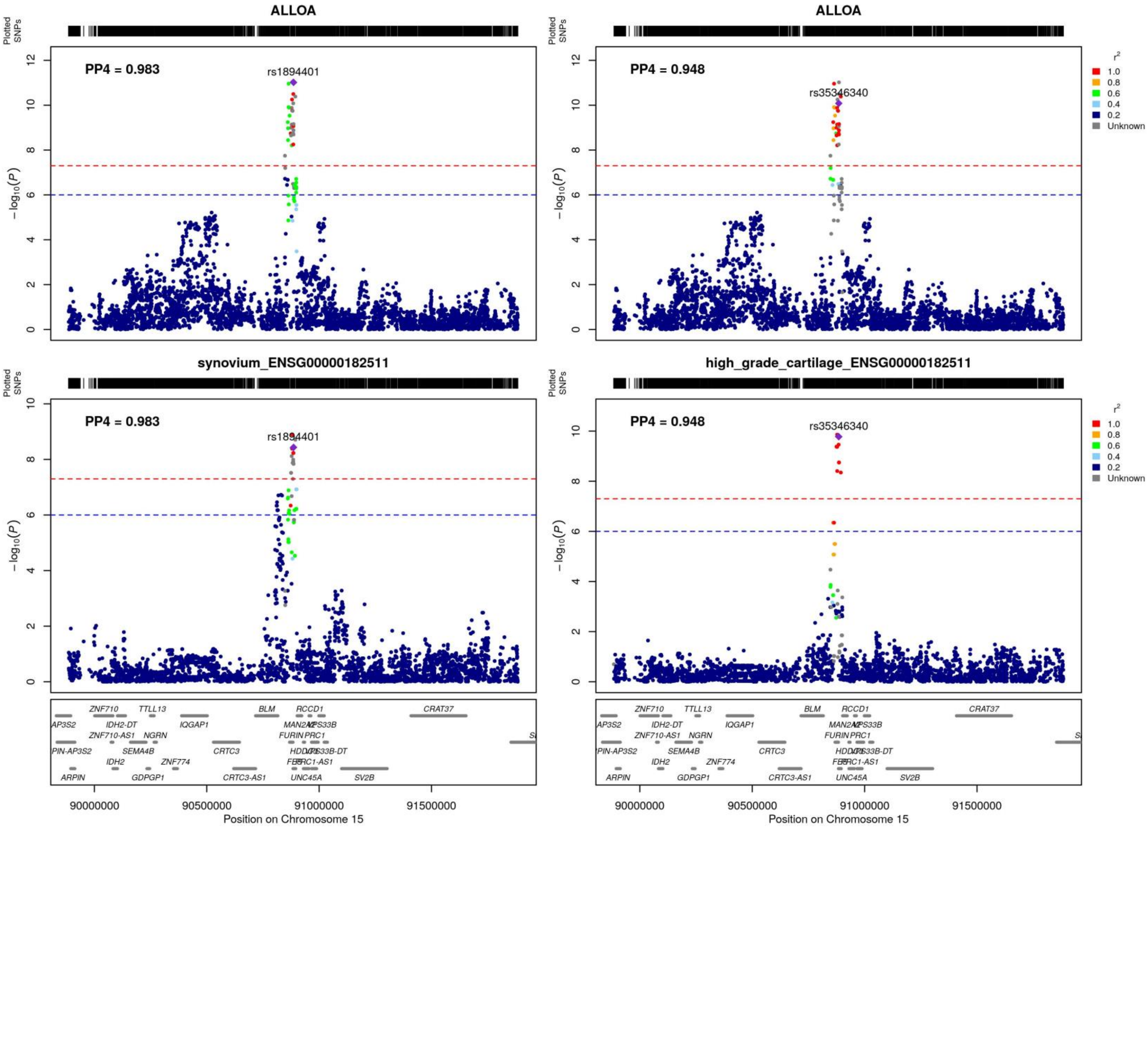

ALLOA
PP4 = 0.983
rs1894401
ALLOA
PP4 = 0.948
rs35346340
synovium_ENSG00000182511
PP4 = 0.983
rs1894401
high_grade_cartilage_ENSG00000182511
PP4 = 0.948
rs35346340
Plotted SNPs
$-\log_{10}(P)$
r²
1.0
0.8
0.6
0.4
0.2
Unknown
Position on Chromosome 15
90000000
90500000
91000000
91500000

**Figure S12**

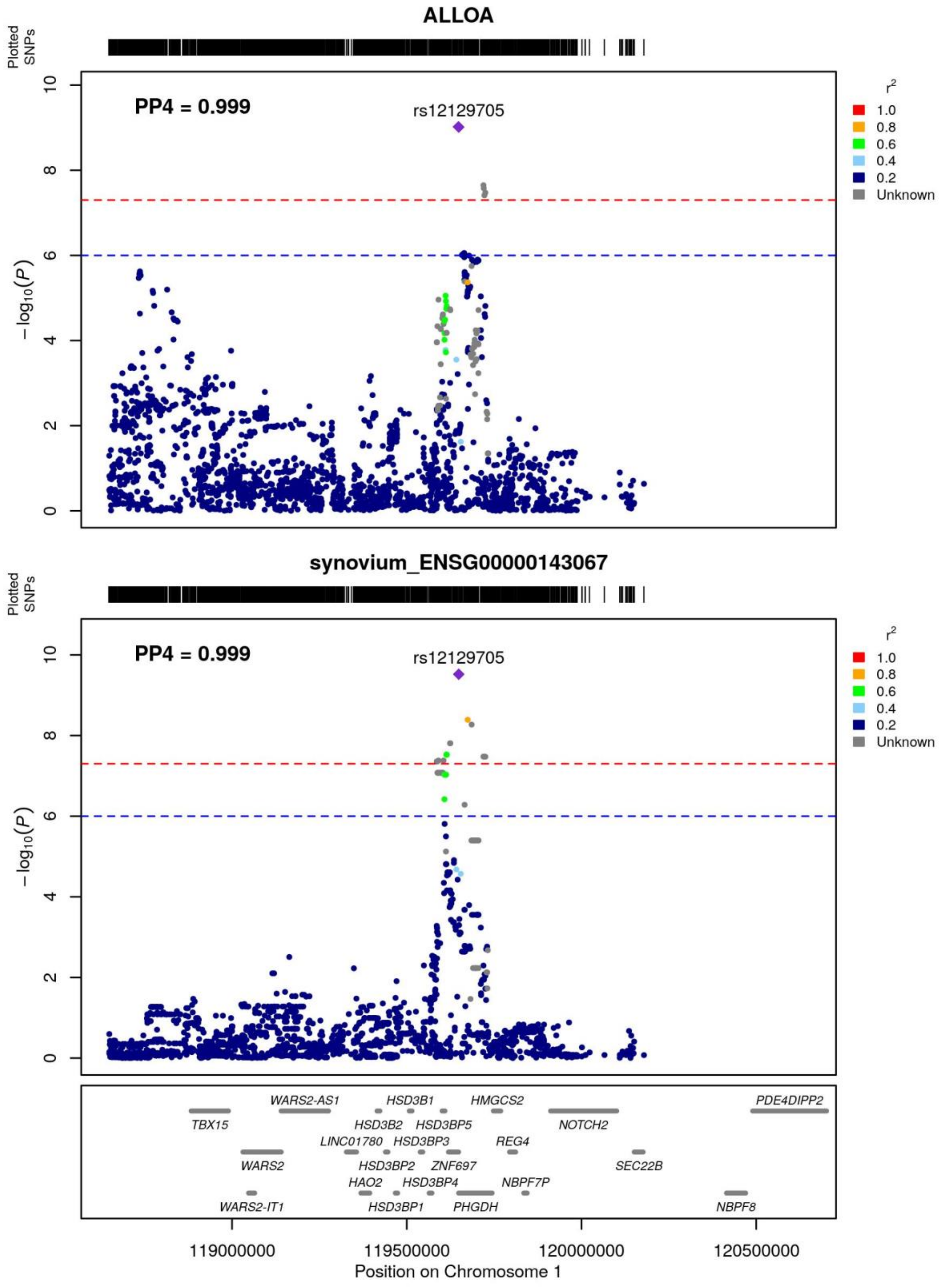
ALLOA
Plotted SNPs
PP4 = 0.999
rs12129705
−log10(P)
r2
1.0
0.8
0.6
0.4
0.2
Unknown
synovium_ENSG00000143067
PP4 = 0.999
rs12129705
TBX15
WARS2-AS1
WARS2
WARS2-IT1
LINC01780
HAO2
HSD3B2
HSD3B1
HSD3BP2
HSD3BP1
HSD3BP5
HSD3BP3
HSD3BP4
ZNF697
PHGDH
HMGCS2
REG4
NBPF7P
NOTCH2
SEC22B
NBPF8
PDE4DIPP2
119000000
119500000
120000000
120500000
Position on Chromosome 1

### Figure S13

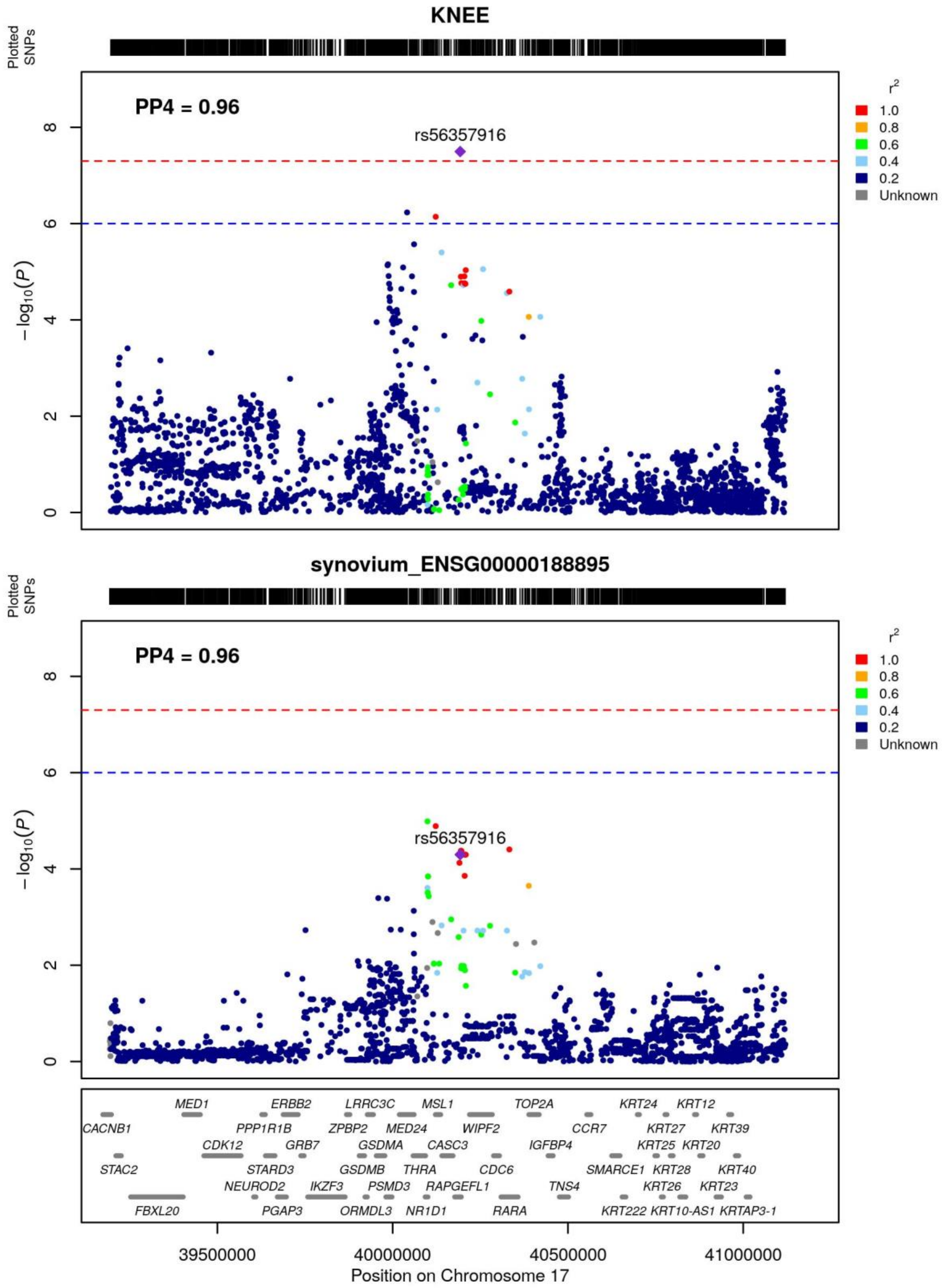

KNEE
Plotted SNPs
PP4 = 0.96
rs56357916
$-\log_{10}(P)$
$r^2$
1.0
0.8
0.6
0.4
0.2
Unknown
synovium_ENSG00000188895
Plotted SNPs
PP4 = 0.96
rs56357916
$-\log_{10}(P)$
$r^2$
1.0
0.8
0.6
0.4
0.2
Unknown
MED1 ERBB2 LRRC3C MSL1 TOP2A KRT24 KRT12
CACNB1 PPP1R1B ZPBP2 MED24 WIPF2 CCR7 KRT27 KRT39
CDK12 GRB7 GSDMA CASC3 IGFBP4 KRT25 KRT20
STAC2 STARD3 GSDMB THRA CDC6 SMARCE1 KRT28 KRT40
NEUROD2 IKZF3 PSMD3 RAPGEFL1 TNS4 KRT26 KRT23
FBXL20 PGAP3 ORMDL3 NR1D1 RARA KRT222 KRT10-AS1 KRTAP3-1
39500000 40000000 40500000 41000000
Position on Chromosome 17
